# Unexplained infertility is frequently caused by defective CatSper function preventing sperm from penetrating the egg coat

**DOI:** 10.1101/2023.03.23.23286813

**Authors:** Samuel Young, Christian Schiffer, Alice Wagner, Jannika Patz, Anton Potapenko, Leonie Herrmann, Verena Nordhoff, Tim Pock, Claudia Krallmann, Birgit Stallmeyer, Albrecht Röpke, Michelina Kierzek, Cristina Biagioni, Tao Wang, Lars Haalck, Dirk Deuster, Jan N Hansen, Dagmar Wachten, Benjamin Risse, Hermann M Behre, Stefan Schlatt, Sabine Kliesch, Frank Tüttelmann, Christoph Brenker, Timo Strünker

## Abstract

The infertility of many couples seems to rest on an enigmatic dysfunction of the men’s sperm, rendering early diagnosis and evidence-based treatment by medically assisted reproduction impossible. Using a novel laboratory test, we assessed the function of the flagellar Ca^2+^ channel CatSper in sperm of almost 2,300 men undergoing a fertility workup. Thereby, we identified a group of men with mutations in *CATSPER* genes affecting the function of the channel. Although standard semen and computer-assisted sperm analysis were unremarkable, the couples required intracytoplasmic sperm injection (ICSI) to conceive a child. We show that their seemingly unexplained infertility and need for ICSI is, in fact, due to the failure of CatSper-deficient human sperm to hyperactivate and penetrate the egg coat. In summary, our study reveals that defective CatSper function represents the most common cause of unexplained male-factor infertility known thus far and that CatSper-related infertility can readily be diagnosed, enabling evidence-based treatment.

## Introduction

Infertility is a multifactorial disease affecting about 50 million couples worldwide; in half of the cases, a male factor contributes to or is solely responsible for the failure to conceive (Agarwal *et al*., 2015). Male-factor infertility is often attributed to an insufficient sperm count (oligo-, crypto-, or azoospermia), impaired sperm motility (asthenozoospermia), and/or abnormal sperm morphology (teratozoospermia). However, for about one third of infertile men, semen parameters are within reference limits (normozoospermia) (Tüttelmann *et al*., 2018). This so called ‘unexplained male infertility’ seems to rest on elusive functional defects of the sperm that are undetectable by standard semen analysis. Moreover, if the fertility workup of the female partner is also unremarkable, affected couples present with ‘unexplained couple infertility’, precluding early diagnosis of the infertility and evidence-based treatment by medically assisted reproduction (MAR) (Barratt *et al*., 2011). As a result, affected couples experience failed MAR attempts, which is not only time consuming and costly, but exposes primarily the female partner to medical risks.

Most sperm functions, e.g., the swimming behavior and acrosomal exocytosis (Alvarez *et al*., 2014; Publicover *et al*., 2008), are controlled by changes in the intracellular Ca^2+^ concentration ([Ca^2+^]_i_). In many species including mammals, [Ca^2+^]_i_ is controlled by the sperm-specific voltage- and alkaline-activated Ca^2+^ channel CatSper (Brown *et al*., 2019; Lishko & Kirichok, 2010; Loux *et al*., 2013; Navarro *et al*., 2008; Quill *et al*., 2001; Rahban & Nef, 2020; Ren *et al*., 2001; Seifert *et al*., 2015; Wang *et al*., 2021a). Mammalian CatSper comprises four homologous pore-forming subunits (CatSper 1-4) (Lin *et al*., 2021; Navarro *et al*., 2008; Wang *et al*., 2021a) and a large number of auxiliary subunits (CatSper β, γ, δ, ɛ, ζ, η, τ, Trim69, Slco6c1, Efcab9, Tmem249 and, perhaps, Cdc42) (Chung *et al*., 2011, 2017; Hwang *et al*., 2019, 2022; Lin *et al*., 2021; Liu *et al*., 2007; Luque *et al*., 2021; Wang *et al*., 2009) for gene/protein nomenclature, see Supplementary Table 1. In mice, targeted ablation of genes encoding CatSper subunits leads to defective channel function, but does not affect sperm number, morphology, or basal motility (Carlson *et al*., 2003, 2005; Chung *et al*., 2011, 2017; Liu *et al*., 2007; Qi *et al*., 2007; Quill *et al*., 2001; Wang *et al*., 2009; Zeng *et al*., 2013). Yet, mouse sperm lacking functional CatSper are unable to switch to so-called hyperactivated motility (Carlson *et al*., 2003; Chung *et al*., 2011, 2014; Ho *et al*., 2009; Jin *et al*., 2007; Qi *et al*., 2007; Quill *et al*., 2003) and, therefore, fail to traverse the oviduct and penetrate the egg coat (Ho *et al*., 2009; Jin *et al*., 2007; Ren *et al*., 2001), resulting in male infertility (reviewed by Wang *et al*., 2021a). Similarly, impaired rather than total loss of CatSper function also affects hyperactivation, resulting in sub- or infertile male mice (Chung *et al*., 2017; Hwang *et al*., 2019, 2022).

These results in mice prompted studies on the role of CatSper in human infertility. Indeed, genetic analysis of consanguineous families with clustering of male-factor infertility identified several men with nonsense variants and homozygous deletions of the *CATSPER1* or *CATSPER2* genes, respectively (Avenarius *et al*., 2009; Avidan *et al*., 2003; Hildebrand *et al*., 2010; Smith *et al*., 2013; Zhang *et al*., 2007). Surprisingly, these men were reported as (oligo)asthenoteratozoospermic, suggesting that in humans, as opposed to mice, CatSper is required for proper sperm production. Of note, in the men lacking *CATSPER2*, the contiguous *STRC* gene encoding stereocilin was also homozygously deleted, which causes mild-to-moderate hearing loss (Verpy *et al*., 2001; Vona *et al*., 2015; Yokota *et al*., 2019). The contiguous deletion of *CATSPER2* and *STRC* was, thus, termed ‘deafness-infertility syndrome’, DIS (OMIM #611102) (Zhang *et al*., 2007).

More recently, the concept that CatSper is required for human spermatogenesis has been challenged by the identification of three men with variants in variants in *CATSPERE* (Brown *et al*., 2018; Williams *et al*., 2015), *CATSPER2* (Luo *et al*., 2019), or *CATSPER3* (Wang *et al*., 2021b), causing loss of channel function. These men were also infertile, but normozoospermic, indicative of defective function rather than production of sperm. Supporting this notion, their sperm also failed to fertilize the egg upon *in vitro* fertilization (IVF), whereas ICSI was successful. Analysis of the phenotype of CatSper-deficient sperm from these three men yielded, however, partly conflicting results or was rather incomplete: For example, hyperactivated sperm motility was abolished (Luo *et al*., 2019), seemingly unaffected (Williams *et al*., 2015), or not even assessed (Wang *et al*., 2021b).

To summarize, despite the discovery of CatSper more than two decades ago, systematic studies on CatSper-deficient men are lacking and results on the reproductive phenotype of those few men identified are conflicting. Accordingly, the role of CatSper in spermatogenesis, sperm function, and infertility in humans has remained ill-defined and controversial.

To solve these long-standing questions, we systematically assessed the function of CatSper in sperm of men undergoing a fertility workup, using a simple motility-based ‘CatSper-Activity-Test’. Thereby, we identified eight men with loss of CatSper function homozygously deleted for *CATSPER2* and one man with impaired channel function featuring bi-allelic variants of *CATSPERE*. CatSper-deficient men presented with unexplained couple infertility involving failure of MAR via ovulation induction (OI), intrauterine insemination (IUI), and/or IVF. By comprehensive microfluidics- and optochemistry-aided motility and flagellar-beat analyses, we revealed the functional deficits of CatSper-deficient human sperm underlying the infertility, failure of OI/IUI/IVF, and need of ICSI. Altogether, we demonstrate that defective CatSper function leads to a similar infertility phenotype in mice and men. Moreover, with an estimated prevalence of about 2.3%, it represents the most common male factor underlying unexplained couple infertility know thus far. Finally, we show that a simple motility-based test can serve as a tool for diagnosis of CatSper-related infertility and evidence-based selection of the technique for medically assisted reproduction.

## Materials and Methods

### Study subjects and ethical approval

The study involved men that underwent semen analysis according to WHO standard procedures (WHO 2010) at the Department of Clinical and Surgical Andrology of the Centre of Reproductive Medicine and Andrology, Münster, as well as healthy volunteers (donors). Clinical data routinely documented in our clinical database *Androbase* (Tüttelmann *et al*., 2006) were used for patient phenotyping (see Table 1 and Supplementary Fig. 4). All participants gave written informed consent according to the protocols approved by the Ethics Committee of the Ärztekammer Westfalen-Lippe and the Medical Faculty Münster (4INie, 2021-402-f-S, and 2010-578-f-S) and according to the Declaration of Helsinki. Genetic analyses were performed within the Male Reproductive Genomics (MERGE) study (Wyrwoll *et al*., 2020). The MERGE subject IDs of the *CATSPER2* and *CATSPERE* patients are: C1 = M1, C2 = M1B1, C3 = M1755, C4 = M1782, C5 = M2107, C6 = M2157, C7 = M2108, C8 = M2212, C9 = M2439.

**Table1.** Semen-analysis parameters and history of medically assisted reproduction of the eight *CATSPER2^-/-^* patients (C1-C8) and the *CATSPERE* ^c.536G>A/c.2394-2399del^ patient (C9)

| Semen Parameters | Reference Values (WHO) | Patient |  |  |  |  |  |  |  |  |
| --- | --- | --- | --- | --- | --- | --- | --- | --- | --- | --- |
|  |  | C1 | C2 | C3 | C4 | C5 | C6 | C7 | C8 | C9 |
| Semen volume (ml) | ≥1.5 | 3.0 | 3.4 | 3.1 | 3.5 | 2.0 | 2.7 | 4.4 | 2.9 | 2.2 |
| pH | ≥7.2 | 7.7 | 8.1 | 7.9 | 8.1 | 8.9 | 7.9 | 7.9 | 8.3 | 8.1 |
| Total sperm count (10<sup>6</sup> per ejaculate) | ≥39 | 440 | 337 | 94 | 90 | 158 | 157 | 224 | 48 | 65 |
| Sperm conc. (10<sup>6</sup> per ml) | ≥15 | 147 | 99 | 30 | 26 | 79 | 58 | 51 | 16 | 30 |
| Total motility (PR + NP, %) | ≥40 | 54 | 47 | 61 | 64 | 63 | 61 | 61 | 56 | 56 |
| Progressive motility (PR, %) | ≥32 | 48 | 40 | 54 | 57 | 56 | 55 | 55 | 50 | 50 |
| Vitality (%) | ≥58 | 75 | 52 | 85 | 72 | 79 | 80 | 74 | 88 | 84 |
| Normal morphology (%) | ≥4 | 7 | 6 | 4 | 4 | 5 | 4 | 6 | 2 | 1 |
| **Success of medically assisted reproduction** |  |  |  |  |  |  |  |  |  |  |
| Ovulation induction (OI) |  | -- | -- | No | -- | -- | No | -- | -- | -- |
| Intrauterine insemination (IUI) |  | -- | -- | -- | No | No | -- | -- | -- | -- |
| In-vitro fertilization (IVF) |  | No | No | No | -- | No | -- | -- | -- | -- |
| Intracytoplasmic sperm injection (ICSI) |  | Yes | Yes | Yes | Yes | Yes | Yes | -- | Yes | -- |
| Live births (thus far) |  | Yes | Yes | -- | Yes | -- | Yes | -- | -- | -- |

### Reagents

Chemicals were purchased from AppliChem (Darmstadt, Germany), Carl Roth (Karlsruhe, Germany), Cayman Chemical (Ann Arbor, USA), Fisher Scientific (Schwerte, Germany), Honeywell Fluka (Charlotte, USA), Sigma-Aldrich (Steinheim, Germany), ThermoFischer Scientific (Waltham, USA), and Tocris Bioscience (Bristol, UK). Human serum albumin (HSA) was purchased from Irvine Scientific (Santa Ana, USA).

### Sperm preparation and buffer conditions

Semen samples provided by donors and patients for research purposes were produced by masturbation and ejaculated into plastic containers. Ejaculates were allowed to liquefy at 37 °C for 30–60 min. Motile sperm were isolated from the ejaculates by a “swim-up” procedure: In a 50-ml conical centrifuge tube, aliquots of 0.5–1 ml ejaculate were layered under 4 ml of human tubal fluid (HTF) buffer containing (in mM): 97.8 NaCl, 4.69 KCl, 0.2 MgSO_4_, 0.37 KH_2_PO_4_, 2.04 CaCl_2_, 0.33 Na-pyruvate, 21.4 lactic acid, 2.78 glucose, 21 HEPES, and 4 NaHCO_3_; adjusted between pH 7.3–7.4 with NaOH. Alternatively, the liquefied semen was diluted 1:10 with HTF, and sperm, somatic cells, and cell debris were pelleted by centrifugation (700 x g, 20 min, 37 °C). The sediment was then resuspended in the same volume of HTF in 50-ml centrifuge tubes. Aliquots of 5 ml of this suspension were pelleted in the 50-ml tubes using a centrifuge with a fixed-angle rotor (700 x g, 5 min, room temperature). In both cases, motile sperm were allowed to swim out of the sediment for at least 60 minutes at 37 °C, collected with the supernatant, and washed twice with HTF (700 x g, 20 min, 37 °C). After the second centrifugation, sperm were resuspended in HTF supplemented with 3 mg/ml HSA (dubbed HTF^+^), or in capacitating HTF^++^ containing (in mM): 72.8 NaCl, 4.69 KCl, 0.2 MgSO_4_, 0.37 KH_2_PO_4_, 2.04 CaCl_2_, 0.33 Na-pyruvate, 21.4 lactic acid, 2.78 glucose, 25 NaHCO_3_, and 21 HEPES, pH 7.35 (adjusted with NaOH), and supplemented with 3 mg/ml HSA. The sperm density was determined using a Neubauer counting chamber and adjusted to 1 x 10^7^ sperm/ml. Sperm were incubated at 37 °C under capacitating conditions in HTF^++^ for at least 2 and 3 h for acrosome-reaction assays and motility analysis, respectively.

For motility experiments designed to study non-capacitated and capacitated sperm side-by-side, an aliquot of the ejaculate of donors or *CATSPER2^-/-^*patients was subjected to swim-up and ensuing washing procedures in bicarbonate-free HTF (HTF^0BC^) containing (in mM): 97.8 NaCl, 4.69 KCl, 0.2 MgSO_4_, 0.37 KH_2_PO_4_, 2.04 CaCl_2_, 0.33 Na-pyruvate, 21.4 lactic acid, 2.78 glucose, and 21 HEPES, pH 7.35 (adjusted with NaOH) without HSA, representing non-capacitating conditions.

### Analysis of sperm-motility decay in Ca^2+^-free buffer

Swim-up sperm in HTF^+^ were diluted tenfold in HTF^+^ or Ca^2+^-free HTF (HTF^0Ca^; [Ca^2+^]_o_ < 20 nM), containing (in mM): 91.8 NaCl, 4.69 KCl, 0.2 MgSO4, 0.37 KH2PO4, 5 EGTA, 0.33 Na-pyruvate, 21.4 lactic acid, 2.78 glucose, 25 NaHCO3, and 21 HEPES, adjusted to pH 7.35 with NaOH, supplemented with 3 mg/ml HSA. The fraction of motile sperm was monitored over time. The time constant, τ, of the motility decay of sperm in HTF^0Ca^ was derived by nonlinear regression analysis using the one-phase decay equation: *Y* = (*Y*_0_ − *plateau*) ∗ *e*^(−*X*·(1/τ))^ + *plateau*, where Y is the fraction of motile cells (%), Y_0_ is the initial fraction of motile cells (constrained to 100%), Plateau is the fraction of motile cells at infinite time (constrained to 0%), and x is the time. To study the role of CatSper in the HTF^0Ca^-induced motility decay, the buffer was fortified with progesterone or RU1968.

### CatSper-Activity-Test

The CatSper-Activity-Test was performed on men undergoing a standard semen analysis in the course of a fertility workup at our centre, provided that the ejaculate featured a total sperm count ≥ 5 × 10^6^ cells and ≥ 10% total motile sperm. We also performed the test on ejaculates from donors. A 20-µl aliquot of the ejaculate was diluted tenfold in HTF^+^ (Buffer A) and HTF^0Ca^ containing 10 µM progesterone (Buffer B) or Buffer B including 5 mM EDTA. To derive the “CatSper-Activity-Index” (CAI), the fraction of motile sperm in Buffer A and Buffer B was determined after 15 or 30 minutes and in Buffer B including EDTA after 30 or 60 minutes according to the following equation: 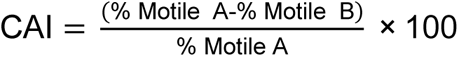.

### Measurement of changes in [Ca^2+^]_i_ in sperm populations

Changes in [Ca^2+^]_i_ were measured in swim-up sperm (in HTF^+^) loaded with the fluorescent Ca^2+^ indicator Fluo-4-AM in 384 multi-well plates in a fluorescence plate reader (Fluostar Omega, BMG Labtech; Ortenberg, Germany) at 30 °C as described before (Brenker, *et al*., 2018a; Schiffer *et al*., 2014; Strünker *et al*., 2011). Briefly, sperm were loaded with Fluo-4-AM (5 µM, 20 min) at 37 °C in the presence of Pluronic F-127 (0.05% w/v). After incubation, excess dye was removed by centrifugation (700 x g, 5 min, room temperature). Sperm were resuspended in HTF at a density of 5 × 10^6^ cells/ml. Each well was filled with 54 µl of the sperm suspension; the fluorophore was excited at 480 nm (Fluo-4), and fluorescence emission was recorded at 520 nm. Changes in Fluo-4 fluorescence were converted to changes in ΔF/F_0_ (%), indicating the percentage change in fluorescence (ΔF) with respect to the mean basal fluorescence (F_0_) before application of buffer or stimuli, respectively. The changes in ΔF/F_0_ (%) evoked by progesterone, PGE1, or NH_4_Cl were normalized to the maximal increase in ΔF/F_0_ (%) evoked by ionomycin to correct for inter-experimental variations in maximal signal amplitudes.

### Single-cell Ca^2^ fluorimetry

Slides (10-well CELLview slide, Greiner Bio-One; Frickenhausen, Germany) were prepared by coating the wells with poly-L-lysine by incubating the empty wells for 60 min at 37 °C in a 1:10 aqueous mixture of poly-L-lysine solution (1 mg/ml) and sodium borate buffer (1 M, pH 8.5). Treated wells were washed with HTF^+^ buffer and left to dry. Swim-up sperm in HTF^+^ were loaded with Fluo-4-AM (5 µM, 60 min) at 37 °C in the presence of Pluronic F-127 (0.05% w/v). After incubation, excess dye was removed by centrifugation (700 x g, 5 min, room temperature). Sperm pellets were resuspended in HTF^+^ to a density of 5 × 10^6^ cells/ml, aliquoted (200 µl) into the prepared wells, and placed in an incubator (37 °C, 10% CO_2_) for one hour to allow sperm to settle. Wells were then washed with HTF^+^ and placed under an inverted microscope (IX73, Olympus; Hamburg, Germany) with a condenser (IX2-LWUCD, Olympus; Hamburg, Germany) and a custom-made dark-field filter, housed in an incubator with circulating air heated to 37 °C, and equipped with a 20× objective (UPLFLN20XPH, Olympus; Hamburg, Germany). Wells were illuminated with an LED (475/35 nm, Thorlabs; Lübeck, Germany) and image sequences were recorded at one frame per second (fps) for up to an hour using the sCMOS camera (Zyla 4.2 Plus, Andor Technology; Belfast, UK). After obtaining a fluorescence baseline, either 22 µl of progesterone (3 µM final) or HTF^+^ (control) was added to the well followed by 24.4 µl of ionomycin (1 µM final) as a positive control. Image sequences were analyzed using ImageJ (NIH; Bethesda, USA) and the change in fluorescence intensity (ΔF/F_0_) of selected cells was normalized to the corresponding ionomycin signal.

### Electrophysiology

We recorded from swim-up sperm in the whole-cell configuration as described before (Strünker *et al*., 2011). Seals between pipette and sperm were formed either at the cytoplasmic droplet or in the neck region in standard extracellular solution (HS) containing (in mM): 135 NaCl, 5 KCl, 1 MgSO_4_, 2 CaCl_2_, 5 glucose, 1 Na-pyruvate, 10 lactic acid, and 20 HEPES, adjusted to pH 7.4 with NaOH. Monovalent currents were recorded in a sodium-based divalent-free solution (NaDVF) containing (in mM): 140 NaCl, 40 HEPES, and 1 EGTA, adjusted to pH 7.4 with NaOH; the pipette (10–15 MΩ) solution contained (in mM): 130 Cs-aspartate, 5 CsCl, 50 HEPES, and 5 EGTA, adjusted to pH 7.3 with CsOH. Data were not corrected for liquid junction potentials.

### Genetics Analyses

Genomic DNA was extracted via local standard methods from peripheral blood or buccal swabs obtained from the patients/family members.

#### Single nucleotide-polymorphism (SNP) array

Genetic workup started with SNP-array analysis using the Infinium CytoSNP-850K v1.2 BeadChip Kit (Illumina; San Diego, USA) according to the manufacturer’s protocol. The BeadChips were scanned using the NextSeq 550 System (Illumina; San Diego, USA). Data was analyzed with BlueFuse Multi v4.5 (Illumina; San Diego, USA).

#### Multiple ligation-dependent probe amplification (MLPA)

To determine the extent of the deletions on chromosome 15 identified in patients C1-C8 via the SNP array, the SALSA MLPA Probemix P461-A1 DIS (MRC Holland; Amsterdam, The Netherlands) kit was used. This MLPA is designed to analyze subjects with sensorineural hearing loss and, therefore, specifically detects deletions or duplications in the genes *STRC*, *CATSPER2* and *OTOA*. Data was analyzed with Coffalyser.Net analysis software (MRC Holland; Amsterdam, The Netherlands).

#### Exome sequencing and analysis

Because patient C9 did not feature a deletion of *CATSPER2*, we sequenced his exome. After enrichment with Twist Bioscience’s Human Core Exome kit, sequencing was conducted on the Illumina NovaSeq 6000® system using the TruSeq SBS Kit v3 - HS (200 cycles). The bioinformatic analyses have been described in detail elsewhere (Wyrwoll *et al*., 2022). In brief, trimming and alignment of reads (Cutadapt v1.18, BWA Mem v0.7.17) and subsequent calling and annotation of variants (GATK toolkit v3.8, Ensembl Variant Effect Predictor v100) were carried out. Exome data were screened focusing on rare (minor allele frequency [MAF] <1% in the gnomAD database, v2.1.1) bi-allelic variants predicted to affect protein function (stop-gain, frameshift, and splice site variants as well as missense variants with a Combined Annotation Dependent Depletion [CADD] score ≥ 20) in genes encoding subunits or regulatory proteins of the CatSper channel (Supplementary Table 1).

Sanger sequencing. Validation of variants in C9 and segregation analysis in family members were performed by Sanger sequencing. Primer sequences are provided in Supplementary Table 2.

### Standard computer-assisted sperm analysis (CASA)

Custom motility chambers (depth ∼60 µm) were prepared by applying two pieces of adhesive tape to a glass slide approximately one centimeter apart. Samples (30 µl) of diluted ejaculates or swim-up sperm were then placed between the strips of adhesive tape and the assembly covered with a coverslip. For basal motility analysis, dark-field images of ejaculates diluted 1:10 in HTF^+^ were recorded with a CMOS camera (UI-3140CP-M-GL R2, iDS GmbH; Obersulm, Germany) at 80 fps for 1 sec with an upright microscope (BX-40, Olympus; Hamburg, Germany) equipped with a heated (37°C) stage (HT 50, Minitüb; Tiefenbach, Germany) and a 10× objective (UPLANFL 10X, Olympus; Hamburg, Germany), and a 0.5× magnification adapter (U-TV0.5XC-3, Olympus; Hamburg, Germany) (total magnification: 5×). Image stacks were then analyzed in ImageJ (NIH; Bethesda, USA) with an open-source CASA program (Giaretta *et al*., 2017; Wilson-Leedy & Ingermann, 2006) adapted for human sperm.

Hyperactivation of non-capacitated swim-up sperm (1 × 10^6^ cells/ml) prepared in HTF^0BC^ was measured by supplementing HSA (3 mg/ml) only immediately prior to the experiment to prevent attaching of sperm to the surface of the recording chamber. Hyperactivation of capacitated swim-up sperm was studied following incubation of sperm for at least 3 h under capacitating conditions in HTF^++^. The sperm sample was diluted to 1 × 10^6^ cells/ml in HTF^++^ before transferring to the recording chamber. Sperm with a curvilinear velocity (VCL) ≥ 150 µm/s, linearity (LIN) ≤ 0.5, and amplitude of lateral head displacement (ALH) ≥ 7 µm were classified as hyperactive (Mortimer *et al*., 1998).

### Microfluidic rapid-mixing CASA

Progesterone-evoked motility changes in swim-up sperm were measured with the inverted IX73 microscope and incubator setup used for single-cell Ca^2^ fluorimetry, equipped with the 0.5× magnification adapter and 10× objective used for standard CASA analysis. A microfluidic chamber with three inlets (µ-Slide III 3in1, Ibidi; Gräfelfing, Germany) was connected via 3/2-way valves to the following reservoirs: (1) capacitated sperm (5 × 10^6^ cells/ml HTF^++^), (2) HTF^++^, and (3) HTF^++^ containing progesterone (10 µM). The single outlet of the chamber was connected to a syringe pump (World Precision Instruments; Sarasota, USA), which, by setting the 3/2-way valves, pulled fluid from two designated reservoirs (e.g., sperm and HTF^++^ (control) or sperm and HTF^++^ containing progesterone) under laminar flow, priming the chamber for mixing. By abruptly reversing the pump, the laminar fluid layers inside the chamber were turbulently mixed. Time-stamped dark-field images (80 fps) were continuously recorded over 180 seconds with the CMOS camera (80 fps) also used for standard CASA. Three one-second image sequences were extracted from the following time frames for analysis with the ImageJ-based CASA program as described before: 0–15, 16–30, 31–45, 46–60, 61–75, 105–120, and 165–180 seconds. The CASA results of the respective time frames were pooled to calculate the overall percentage of hyperactivated sperm.

### Flagellar-beat analysis of head-tethered sperm

The basal flagellar waveform of head-tethered swim-up sperm was analyzed using the IX73 microscope, condenser, and dark-field filter setup also used for single-cell Ca^2+^ fluorimetry, equipped with a 20× objective (UPLFLN20XPH, Olympus; Hamburg, Germany). Dark-field image sequences (250 fps, 5 sec) of single spermatozoa were recorded using a sCMOS camera (Zyla 4.2 Plus, Andor Technology; Belfast, UK) and analyzed in ImageJ using the software tool ”SpermQ” (Hansen *et al*., 2018); https://github.com/hansenjn/SpermQ. To measure progesterone-evoked motility responses, sperm suspension was fortified with 2 µM caged-progesterone (Kilic *et al*., 2009). Head tethered sperm were recorded (135 fps) 15 seconds before and after photo-release (2 sillumination at 365 nm) of progesterone from the caged-progesterone.

#### Asymmetry Index

The maximal beat amplitude along the arc length of the flagellum both above and below the head-midpiece axis of the sperm was measured to determine the beat envelope. The “asymmetry index” was calculated by taking the ratio of the area under the beat envelope, i.e. area under the curve (AUC), generated by the beat envelope above (↑AUC) and below (↓AUC) the head-midpiece axis, respectively, according to the following equation:

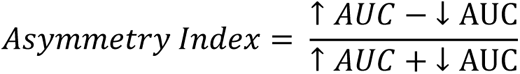

Hence, an asymmetry index of 0 indicates perfect beat symmetry, while a value of 1 indicates complete asymmetry. The asymmetry index was measured before and after the photo-release of progesterone from the caged progesterone.

### Viscous media penetration test

Capillary tubes (0.2 × 4.0 mm) (VitroTubes, VitroCom; Mountain Lakes, USA) were filled by capillary action with HTF^++^ supplemented with 1% w/v methyl cellulose containing either progesterone (3 µM) or vehicle (DMSO) and capped with sealing wax (Glaswarenfabrik Karl Hecht; Sondheim vor der Rhön, Germany). Swim-up sperm were capacitated in HTF^++^ in aliquots of 300 µl at 3 × 10^6^ cells/ml. After adding either progesterone (3 µM) or vehicle (DMSO), the filled capillary tubes with the corresponding content were placed upright in these sperm suspensions, and placed in an incubator (37 °C, 10% CO_2_). After one hour, sperm within a field-of-view corresponding to 600 µm above and below the 2-cm mark inside the capillary were counted, and the fraction of motile sperm in the suspension was determined using the upright BX-40 microscope. The number of cells at the 2-cm mark was normalized to the fraction of motile sperm, correcting for inter-sample variation.

### Acrosome Reaction

The evaluation of the acrosome reaction was performed based on a slightly modified protocol previously described (Rennhack *et al*., 2018; Tamburrino *et al*., 2014). Briefly, capacitated swim-up sperm in HTF^++^ (2 × 10^6^ cells/ml) were incubated with either progesterone (5 µM) or ionomycin (5 µM) for 1 h at 37 °C, or supplemented with the equivalent volume of HTF^++^ (control). Afterwards, sperm were washed by centrifugation (700 × g, 5 min, room temperature) and resuspended in a hypo-osmotic swelling medium (1:10 HTF^++^:ddH_2_O) for 1h at 37°C. After a second washing step, sperm were fixed in 50 μl of ice-cold methanol, transferred on a slide in five aliquots (10 μl per spot), air-dried, and stored at −20 °C. For acrosome staining, the fixed sperm were thawed and incubated with 1 mg/ml FITC-labelled *Arachis hypogaea* (peanut) agglutinin (PNA) lectin (Sigma Aldrich; Steinheim, Germany) in PBS at 4 °C for 20 min in the dark. Slides were washed with PBS and incubated for 10 s with 5 mg/ml DAPI (ThermoFischer; Waltham, USA) in ddH_2_O at room temperature, washed again with PBS, and air dried. Images from the slides were taken using an AXIO Observer microscope (Carl Zeiss; Jena, Germany). For each condition, 200 curled-tail (viable) sperm were considered for analysis. Acrosomal status was assessed manually in a blinded fashion by two independent operators using a custom-designed counting aid software. The results of the two independent assessments were averaged.

### Data analysis

Experiments and data analysis were performed without randomization and blinding, except for the acrosome reaction test. All data is presented as mean ± standard deviation (SD), unless otherwise stated. Statistical analysis was performed using GraphPad Prism 5 (GraphPad Software; San Diego, USA). If the experiment involved two conditions (control and treatment), a two-tailed paired t-test was used. Results from donors and CatSper-deficient patients obtained under the same conditions were also analyzed using two-tailed unpaired t-test. ANOVA was used for experiments involving one parameter measured over time (repeated measures with Dunnett’s multiple comparisons *post hoc* test) or more than one treatment (one-way with Bonferroni’s multiple comparisons *post hoc* test).

## Results

### A simple motility-based test allows to determine the activity of CatSper in human sperm

To elucidate the role of CatSper in male-factor infertility, we set out to develop a simple laboratory test that identifies patients with defective CatSper function. Lowering the extracellular Ca^2+^ concentration ([Ca^2+^]_o_) to nanomolar levels renders mouse (Jin *et al*., 2007) and human sperm (Schiffer *et al*., 2020; Torres-Flores *et al*., 2011) immotile. In CatSper-deficient mouse sperm, the cessation of motility by low [Ca^2+^]_o_ is abolished (Jin *et al*., 2007), demonstrating that it requires functional CatSper. We tested whether this holds true for human sperm. An aliquot of the ejaculate from donors with normal CatSper function was diluted tenfold with control (HTF^+^) or Ca^2+^-free HTF buffer (HTF^0Ca^, [Ca^2+^]_o_ < 20 nM), and the ensuing change in the fraction of motile sperm was monitored over time. In control buffer, the fraction of motile sperm remained constant, whereas in Ca^2+^-free buffer, it decreased exponentially with a time constant (τ) of 27 ± 11 min (mean ± standard deviation (SD), n = 9) (Fig. 1A). In Ca^2+^-free buffer fortified with the CatSper-inhibitor RU1968 (Rennhack *et al*., 2018), the decrease was abolished (Fig. 1A), indicating that it requires functional CatSper. Of note, in human sperm, CatSper is activated by steroids and prostaglandins contained in reproductive fluids (Brenker *et al*., 2018b; Jeschke *et al*., 2021; Lishko *et al*., 2011; Strünker *et al*., 2011), and whereas inhibition of CatSper abolished the motility decrease in Ca^2+^-free buffer, it was accelerated about fourfold upon activation of the channel by progesterone (Fig. 1A). We surmised that the action of low [Ca^2+^]_o_ on human sperm motility can be harnessed to assess the activity of CatSper by an end-point motility test.

**Figure 1:**
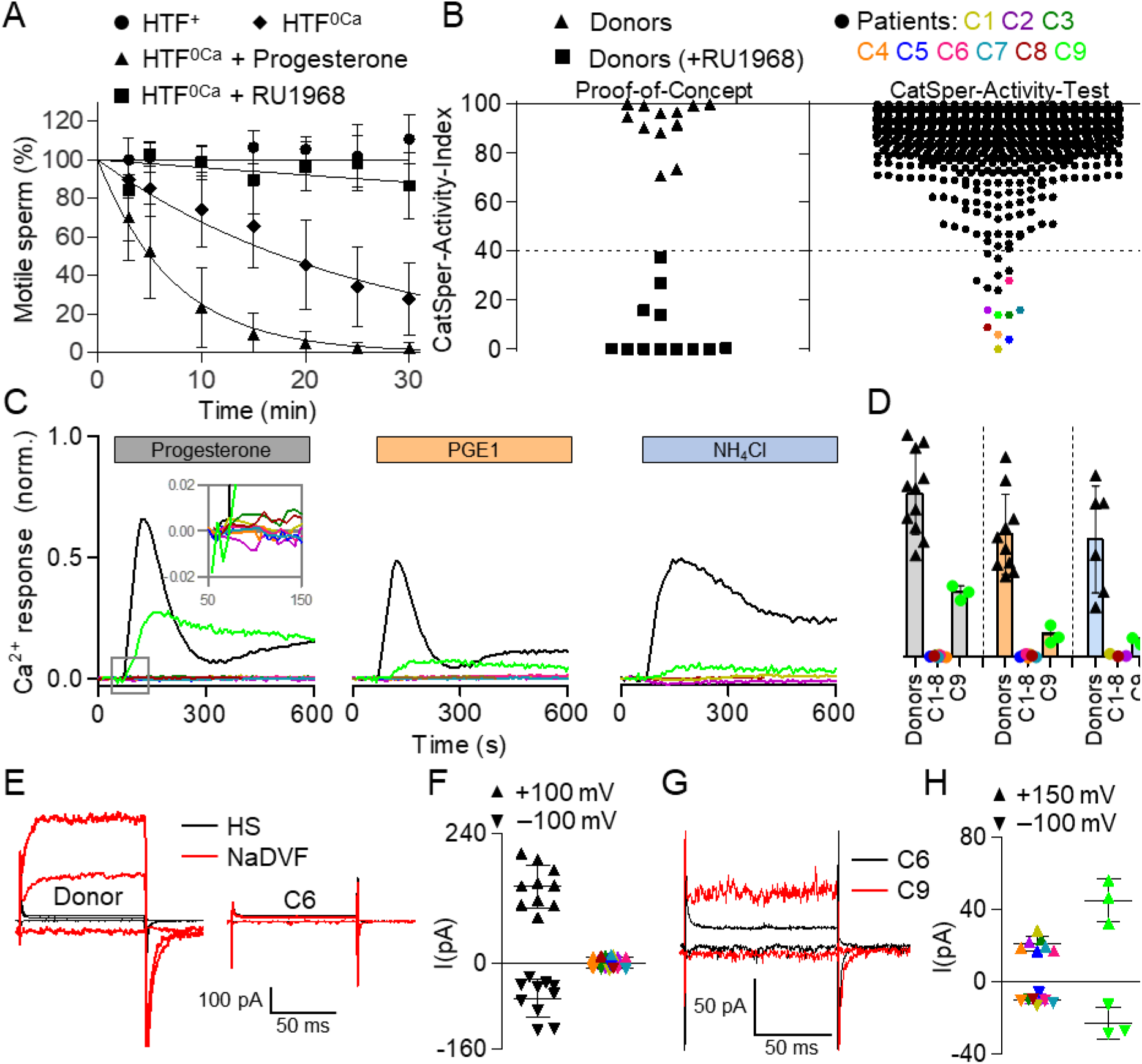
Identification of patients with loss or impaired CatSper function. **(A)** Changes in the fraction of motile sperm (mean ± SD) upon dilution of semen samples from donors in Ca^2+^-free HTF (HTF^0Ca^) (diamonds), HTF^0Ca^ containing progesterone (10 µM) (triangles), or HTF^0Ca^ containing the CatSper-inhibitor RU1968 (15 µM) (squares), relative to the fraction of motile sperm determined upon dilution of the respective semen sample in control HTF^+^ (circles) at t = 0 (set to 100%). An exponential decay curve was fitted to the change in the fraction of motile cells averaged over all replicates. **(B) Left**: CatSper-Activity-Indices (CAI, see explanation in the text) determined 15 minutes after dilution of semen samples from donors in HTF^+^ (Buffer A) and HTF^0Ca^ containing progesterone (triangles) (Buffer B) or Buffer B also containing RU1968 (15 µM) (squares). **Right:** CAI values from semen samples of men undergoing semen analysis (n = 2,286); see explanation in the text as to the dotted line. Patients with confirmed loss or impaired CatSper function are labeled C1-C9 and indicated with a color-coded circle. **(C)** Representative Ca^2+^ signals in sperm from patient C1-C9 (color coded) and a donor (black) evoked by progesterone (3 µM), PGE1 (3 µM), or NH_4_Cl (30 mM) relative to the maximal signal amplitude evoked by ionomycin (3 µM) (set to 1). **(D)** Mean (± SD) maximal Ca^2+^-signal amplitude in sperm from donors (black) and patients C1-C9 (color coded) evoked by progesterone (3 µM) (gray), PGE1 (3 µM) (orange), or NH_4_Cl (30 mm) (blue) relative to that evoked by ionomycin (set to 1). **(E) Left:** Representative whole-cell currents recorded from a donor sperm cell at pH_i_ 7.3 in extracellular solution containing Mg^2+^ and Ca^2+^ (HS) and in Na^+^-based divalent-free bath solution (NaDVF). Currents were evoked by stepping the membrane voltage from -100 to +100 mV and +150 mV from a holding potential of -80 mV. **Right:** Representative currents recorded under the same conditions from a sperm cell from patient C6; the prototypical monovalent CatSper currents in NaVDF are abolished. **(F)** Steady-state current amplitudes at +100 mV and -100 mV in NaDVF recorded from sperm from donors (black) and patients C1-C8 (color coded). **(G)** Representative whole-cell currents recorded from patient C6 (black) and patient C9 (red) at pH_i_ 7.3 in NaDVF. Currents were evoked by stepping the membrane voltage from -100 to +150 mV from a holding potential of -80 mV. **(H)** Steady-state current amplitudes at +150 mV and -100 mV in NaDVF recorded from sperm from patients C1-C8 (color coded) as shown in G (consider the different scales of Y-axes) compared to that recorded in sperm from patient C9 (light green). Replicates shown for patient C9 in (D) and (H) represent results from experiments performed independently on sperm from three different semen samples. Of note, data on Ca^2+^ responses and membrane currents in sperm from patient C1-C5 comprise data from Schiffer *et al*. (2020), reporting on these five patients for the first time, combined with data from additional experiments.

As a proof-of-concept, we diluted the ejaculate from donors with HTF^+^ (Buffer A) and HTF^0Ca^ containing progesterone (Buffer B). After 15 minutes, the fractions of motile sperm in A and B were determined and a ‘CatSper-Activity-Index (CAI)’ was calculated 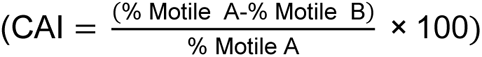, which ranged from 70–100; however, when Buffer B was fortified with RU1968, the CAI decreased to < 40 (Fig. 1B, left), reflecting that inhibition of CatSper rescues sperm motility in Buffer B. These results supported the notion that this motility-based test, which we refer to as the CatSper-Activity-Test, allows to systematically assess the activity of CatSper in human sperm.

### The CatSper-Activity-Test identifies patients with defective CatSper function

We next performed the CatSper-Activity-Test over a period of three years on 2,286 men undergoing semen analysis in the course of a fertility workup. In more than 99%, the CAI was > 40 (Fig. 1B, right), which we regarded as indicative of rather normal CatSper function; prototypical CatSper-mediated Ca^2+^ signals recorded in sperm from a representative sample of 48 of those men supported this assumption (not shown). In 16 men subjected to the CatSper-Activity-Test, the CAI was however similar to that in donors upon inhibition of CatSper, i.e., ≤ 40, indicative of defective CatSper function (Fig. 1B, right). A series of follow-up experiments confirmed that nine of these men – in the following referred to as patients C1–C9 (Fig. 1B, right, color-coded circles) – indeed featured defective CatSper function. Sperm isolated from ejaculates of patients C1–C8 lacked the prototypical Ca^2+^ signals evoked by progesterone-, prostaglandin E1-, or alkaline-activation of CatSper in sperm populations (Fig. 1C, D, colored traces and circles); the Ca^2+^ signal evoked by the Ca^2+^ ionophore ionomycin was however preserved. For patients C1 and C2, the lack of CatSper-mediated Ca^2+^ signals was validated on the single-cell level (Supplementary Fig. 1). Electrophysiological recordings revealed that sperm from patients C1-C8 also lacked CatSper-mediated membrane currents (Fig. 1E, F). Furthermore, in sperm from patient C9 (Fig. 1B, green circle), CatSper-mediated Ca^2+^ signals (Fig. 1C, D, green trace and circles, respectively) were 5- to 10-fold reduced in amplitude and featured an atypical mono-rather than biphasic waveform (Fig. 1C); CatSper-mediated membrane currents were also largely, but not entirely, abolished (Fig. 1G, H). Thus, whereas sperm from patients C1–C8 lacked functional CatSper channels, patient C9 featured a severely impaired CatSper function. These results demonstrate that a fraction of men undergoing a fertility workup suffers from defective CatSper function, which can readily be identified by the CatSper-Activity-Test.

Of note, as detailed in Supplementary Fig. 2, we successively optimized the CatSper-Activity-Test during the course of screening to minimize and, finally, avoid false positive results. Moreover, using ejaculates from donors and CatSper-deficient patients as a tool, we revealed that the test distinguishes best between men with normal versus loss of CatSper function when the fraction of motile sperm is determined 60 min after dilution in the test buffers. However, additional studies are required to determine the test’s sensitivity and specificity using the optimized conditions.

### Defective CatSper function is linked to germline mutations in *CATSPER* genes

Genetic workup elucidated the pathomechanisms underlying the defective CatSper function: Patients C1–C8 shared a homozygous deletion of the *CATSPER2* gene located on chromosome 15 (Fig. 2A, Supplementary Fig. 3, Supplementary Table 3). In patients C1–C5, the deletions were similar on both homologues chromosomes and involved, except for patient C5, also the homozygous deletion of the *CKMT1B* and *STRC* genes (Fig. 2A, Supplementary Fig. 3). In patients C6–C8, the homologous chromosomes exhibited distinct deletions, resulting in heterozygous deletion of *STRC* and *CKMT1B* (Fig. 2A, Supplementary Fig. 3). Of note, patient C1 and C2 are brothers, and their parents as well as another brother are heterozygous carriers of the deletion, whereas their sister is homozygous (Fig 2C).

**Figure 2:**
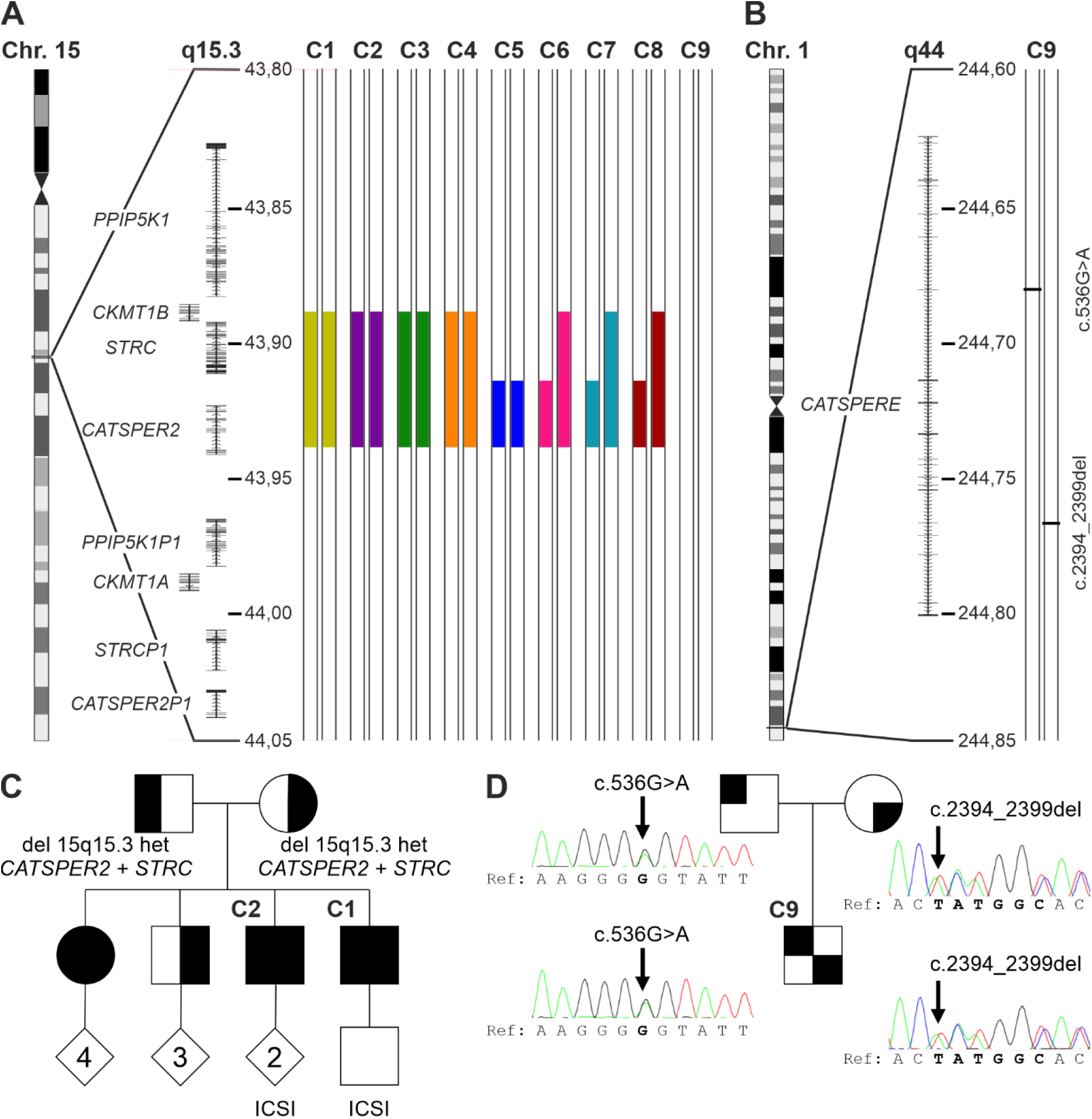
Genetic aberrations identified in CatSper-deficient patients. **(A)** Schematic depiction of chromosome 15, magnified region q15.3 including genes, and the identified deletions (represented as filled color-coded bars) in patients C1-C8, but not patient C9. All positions according to hg19/GRCh37. **(B)** Schematic depiction of chromosome1, magnified region q44 and compound-heterozygous variants (see panel D) (c.536G>A and c.2394_2399del) of *CATSPERE* (NM_001130957.2) identified in patient C9. **(C)** Family pedigree of patients C1 and C2, who are brothers. Their sister is also homozygous for the deletion at 15q15.3, whereas their mother, father, and third brother are heterozygous carriers. **(D)** Family pedigree of patient C9, demonstrating that the father and mother are carriers of the missense variant (c.536G>A) and in-frame deletion (c.2394_2399del), respectively. Of note, in Schiffer *et al*. (2020), we already showed array-CGH data from patients C1-C5, reporting on the deletion of *CATSPER2* in these five patients for the first time.

Patient C9 carried the compound-heterozygous nucleotide variants c.536G>A and c.2394_2399del in the *CATSPERE* gene (NM_001130957.2) located on chromosome 1 (Fig. 2B) inherited from the father and mother, respectively (Fig. 2D). The according to gnomAD novel c.536G>A variant causes a possibly deleterious amino acid substitution p.(Gly179Glu) (CADD score = 25.7). The variant c.2394-2399del causes an in-frame deletion of two amino acids p.Met799_Ala800del, features an allele frequency of 0.001183 (gnomAD), and has already been described in association with CatSper-related male infertility (Brown *et al*., 2018; Williams *et al*., 2015).

In summary, homozygous deletion of *CATSPER2* (*CATSPER2^-/-^*) underlies the loss of CatSper function in patients C1-C8. The severely impaired CatSper function in patient C9 is almost certainly due to the compound-heterozygous variants in *CATSPERE*.

### *CATSPER2^-/-^* patients are normozoospermic, but infertile, and require ICSI to father a child

Next, we assessed the clinical phenotype of the patients. Patients C1-C7 were normozoospermic, i.e., sperm number, motility, and morphology were within reference limits (Table 1, Supplementary Movie 1), demonstrating that deletion of *CATSPER2* and loss of CatSper function do not affect the production of sperm. In patients C8 and C9, sperm number and motility were also within reference limits, but both featured mild teratozoospermia.

Patients C1–C6 and C8 presented with unexplained couple infertility, and in patients C1–C6, not only natural conception, but also medically-assisted reproduction via OI, IUI, and/or IVF failed (Table 1). For patient C5, an IVF/ICSI attempt was documented by video microscopy. Ten minutes after joining the gametes, sperm were already found bound to the oocytes’ surfaces, which after 24 hours, were enveloped by sperm (Fig. 3A, Supplementary Movie 2). However, none of the five oocytes subjected to IVF were fertilized (Fig. 3A), indicating that the sperm failed to penetrate the zona pellucida. Supporting this notion, ICSI performed on 15 oocytes in parallel to the IVF yielded nine embryos (Fig. 3B), corresponding to a fertilization rate of 60%. Similarly, in patients C1–C4, C6, and C8, fertilization was only achieved by ICSI also resulting in live births (Table 1). Finally, patients C7 and C9 were childless, but did not present due to suspected infertility; the identification of the defective CatSper function as well as the underlying genetic aberrations came as incidental findings. Altogether, these results demonstrate that loss of CatSper function results in, according to standard fertility workup, unexplained infertility involving OI, IUI, and IVF failure.

**Figure 3:**
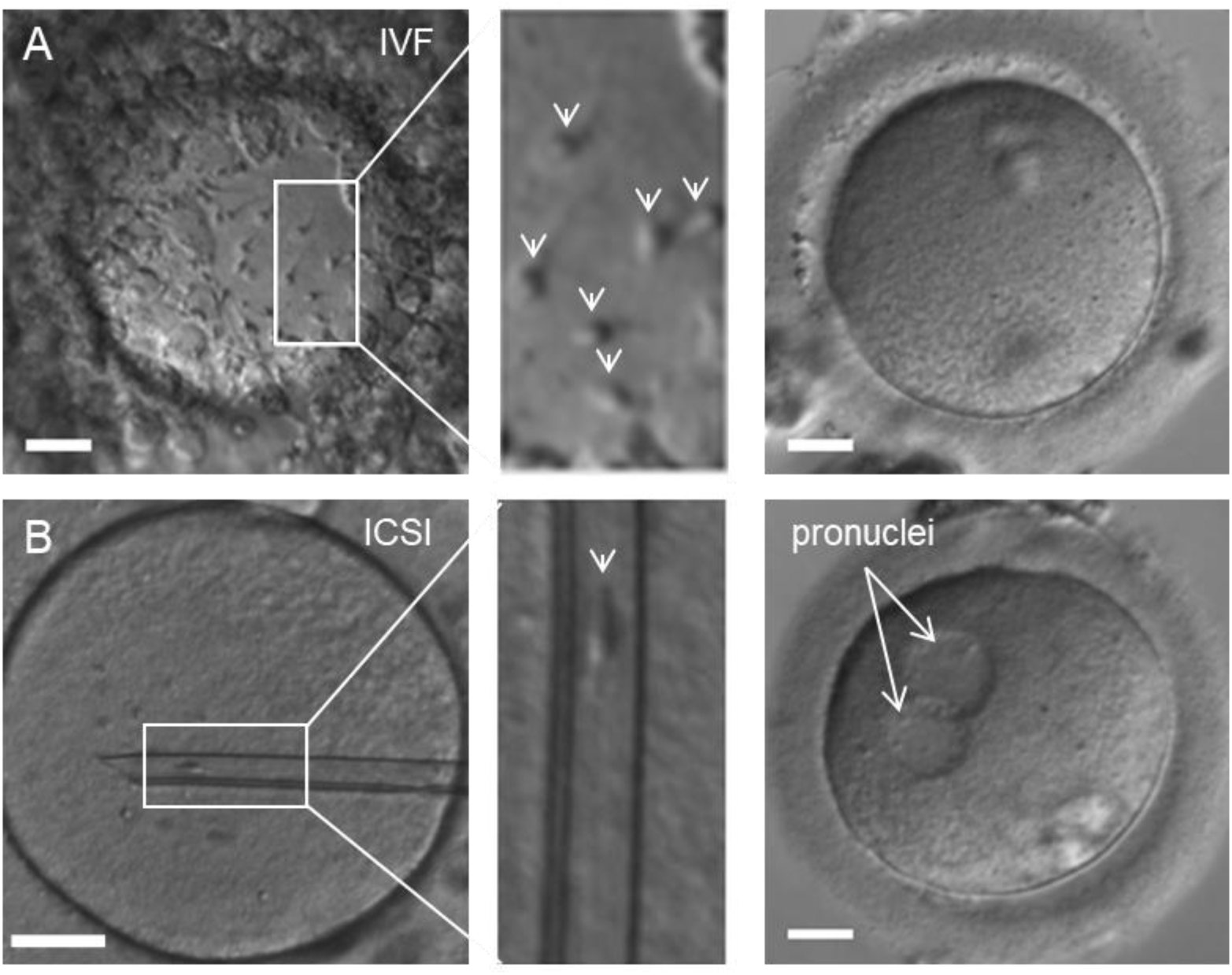
Microscopic documentation of medically-assisted reproduction with *CATSPER2^-/-^* sperm. **(A)** Left: Representative micrograph of an oocyte with *CATSPER2^-/-^* sperm from patient C5 attached to thezona pellucida (indicated by arrows in the zoomed-in image, **middle**), taken after overnight incubation of sperm and oocyte for in-vitro fertilization (IVF). None of the five oocytes subjected to IVF was fertilized (**right**, representative example). **(B) Left:** Representative micrograph of an oocyte with an inserted glass pipette containing a *CATSPER2^-/-^* sperm cell form patient C5 (indicated in the zoomed-in image, **middle**) for intracytoplasmic sperm injection (ICSI). Nine out of the 15 eggs injected with *CATSPER2^-/-^* sperm developed two pronuclei (**right**, representative example) indicating fertilization. Scale bars represent 30 µm.

Stratification of the men enrolled in our study (Supplementary Fig. 4) revealed that 1.2% of normozoospermic men presenting with couple infertility featured a defective CatSper function. Assuming that in half of the infertile couples including a normozoospermic men, the infertility is rather due to a female factor, we can estimate a prevalence of 2.3% for CatSper-related male-factor infertility among couples presenting with unexplained infertility (Supplementary Fig. 4).

We further examined whether patients lacking both *CATSPER2* and *STRC* (*STRC^-/-^*) exhibited sensorineural deafness. Standard audiometry indeed revealed a mild-to-moderate hearing impairment in patients C1–C4 (Supplementary Fig. 6), confirming that they suffer from the deafness-infertility syndrome. Of note, the cause of their hearing impairment was not known to the patients before. This is not surprising: although it is well-known that variants in *STRC* represent the second most common genetic aberration causing mild-to-moderate hearing loss (prevalence ∼4%) (Yokota *et al*., 2019), the gene is still not routinely assessed in the genetic workup of hearing impairment. Therefore, deletions of *STRC* and, thus, also the deafness-infertility syndrome are largely underdiagnosed. Expectedly, in patient C5 with unaffected *STRC* and patient C7 with a heterozygous deletion of *STRC* (*STRC*^+/-^), the audiogram was within normal limits (Supplementary Fig. 6). For *STRC*^-/+^ patients C6 and C8, we did not obtain an audiogram, but neither patient reported impaired hearing.

### Loss of CatSper function affects sperm hyperactivation and migration in viscous media

Next, we set out to unravel the pathomechanism underlying the infertility of *CATSPER2^-/-^* patients and the failure of OI, IUI, and IVF. To this end, we investigated the motility of their sperm both in population and on the single cell level. First, we diluted the ejaculate of donors (control) and patients with HTF^+^ and quantified the basal motility parameters of sperm in population by standard computer-assisted sperm analysis (CASA) (Fig. 4A). The individual motility parameters were largely similar in *CATSPER2^-/-^* and control sperm (Fig. 4B), rendering defective CatSper function rather undetectable by CASA. However, when averaged over all donors and patients, beat-cross frequency (BCF; reflecting the beat frequency) and indicators of the linearity of the swimming path (i.e., LIN, STRC) were slightly enhanced in *CATSPER2^-/-^* sperm (Fig. 4B). Thus, sperm lacking functional CatSper tend to swim with a slightly higher beat frequency and somewhat straighter. Next, we isolated via the swim-up technique non-capacitated, motile sperm from the ejaculate of donors and *CATSPER2^-/-^* patients, tethered them with their head to the surface of a recording chamber, and analyzed the flagellar beat of individual sperm using the SpermQ software (Hansen *et al*., 2018) (Fig. 4C–E). Compared to control sperm, *CATSPER2^-/-^* sperm beat with slightly higher frequency and reduced amplitude (Fig. 4C–E), matching the outcome of CASA. These minor anomalies, however, do not explain the infertility and IUI/IVF failure, but rather demonstrate that CatSper does not play a critical role in the control of basal motility features (Luo *et al*., 2019; Wang *et al*., 2021b; Williams *et al*., 2015). Supporting this notion, in a previous study using motile sperm isolated from the ejaculate of some of the *CATSPER2^-/-^* patients characterized in detail herein, we showed that loss of CatSper function does not affect rotational motion and rheotaxis either (Schiffer *et al*., 2020).

**Figure 4:**
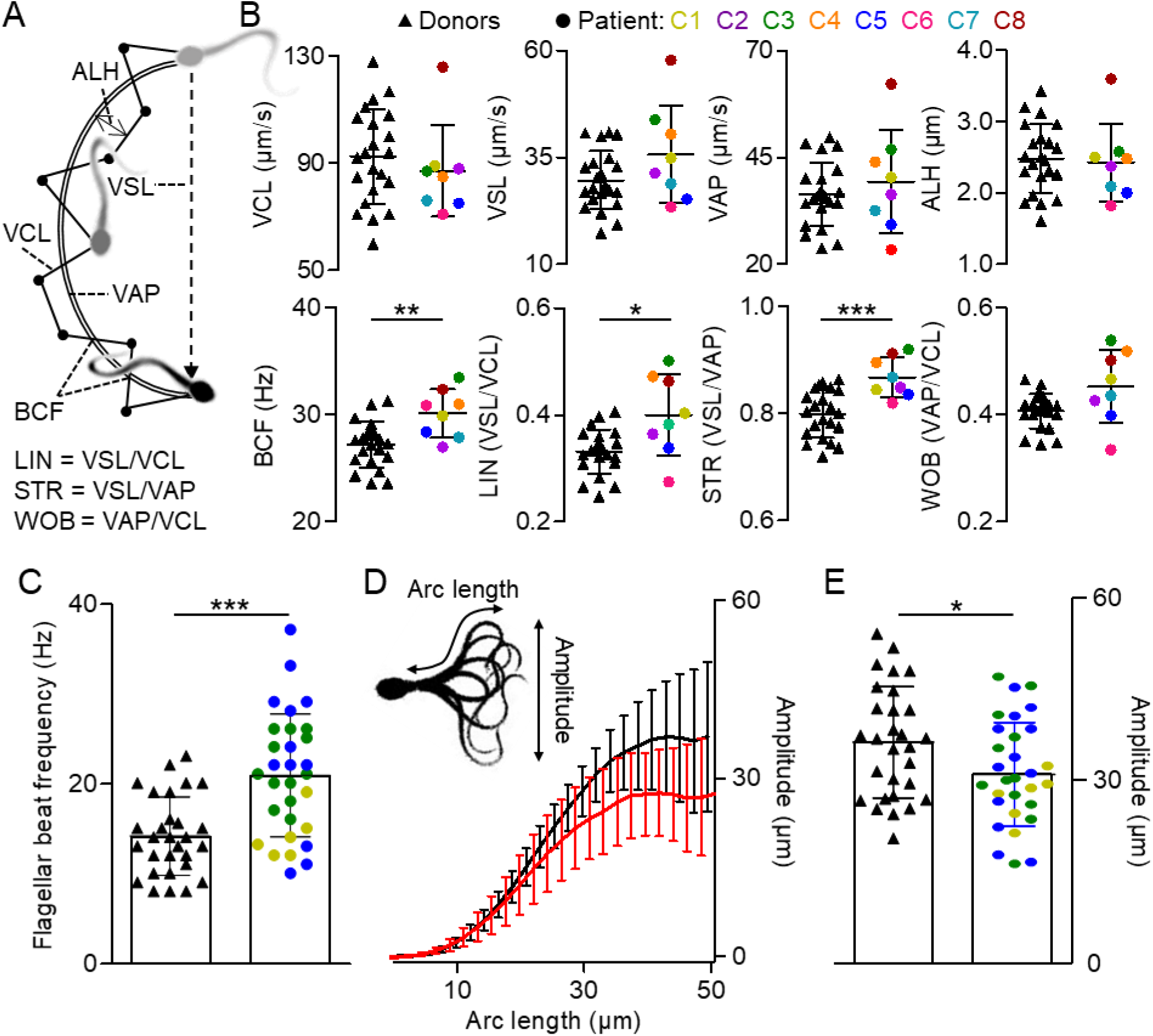
Analysis of basal motility and flagellar beat of control and *CATSPER2^-/-^* sperm. **(A)** Illustration of the swimming path and kinematic parameters of a sperm cell determined by computer-assisted sperm analysis (CASA). Curvilinear velocity (VCL) represents the frame-to-frame track of the sperm head, from which the average-path velocity (VAP) is calculated. The beat-cross frequency (BCF) is the frequency at which the sperm track crosses the VAP path. The amplitude of lateral head displacement (ALH) is the average deviation of the head from the VAP path. The straight-line velocity (VSL) is derived by charting a direct path between the first and last head position in the image sequence. The linearity (LIN), straightness (STR), and wobble (WOB) are indicators for the linearity of the path trajectory. **(B)** Scatter plots (mean ± SD) of kinematic parameters of control (black triangles, n = 22) and *CATSPER2^-/-^* sperm from patients C1-C8 (color-coded circles). **(C)** Scatter plots (mean ± SD) of the flagellar beat frequency of single head-tethered control sperm from donors (black triangles, n = 29 from six experiments) and of sperm from *CATSPER2^-/-^* patients (color-coded circles, n = 30 from three experiments). **(D)** Maximal beat amplitude (mean ± SD) along the arc length of the flagellum of the head-tethered control (black) and *CATSPER2^-/-^* (red) sperm analyzed in (C). **(E)** Scatter plots (mean ± SD) of the maximal beat amplitude of the control (black triangles) and *CATSPER2^-/-^* (color-coded circles) sperm reported on in (C) and (D). *P < 0.05, **P < 0.01, ***P < 0.001, unpaired t-test with Welch’s correction.

However, to penetrate the zona pellucida and fertilize the oocyte, sperm must switch to hyperactivated motility, a powerful flagellar beat characterized by high asymmetry and low frequency resulting in a tortuous swimming trajectory (Suarez, 2008). The role of CatSper in human sperm hyperactivation has remained controversial (Brown *et al*., 2019; Wang *et al*., 2021a). We first determined whether spontaneous hyperactivation that develops during capacitation – a maturation process of sperm inside the female genital tract – is impaired or even abolished in *CATSPER2^-/-^* sperm. To this end, we incubated motile sperm under capacitating conditions in HTF^++^ and determined the fraction of hyperactive sperm by CASA. In donors, the fraction of hyperactive sperm was 12.2 ± 6.1% (n = 17), whereas it was only 0.8 ± 1.0% (n = 10) in *CATSPER2^-/-^* patients (Fig. 5A), demonstrating that loss of CatSper function abolishes spontaneous capacitation-induced hyperactivation (for more details, see Supplementary Fig. 6). We next studied the action of progesterone on the swimming behavior and flagellar beat of capacitated control and *CATSPER2^-/-^* sperm using microfluidics- and optochemistry-aided motility and flagellar-beat analyses. In fact, whether progesterone evokes hyperactivation and whether this can be detected by CASA have been long-standing controversial issues (Baldi *et al*., 2009; Brown *et al*., 2019). Indeed, when determined five minutes after stimulation, progesterone did not significantly increase the fraction of hyperactive sperm (Fig. 5B). Considering the transient nature of progesterone-induced Ca^2+^ signals (see Fig. 1C), we surmised that progesterone-evoked motility responses might also be rather transient. Therefore, we combined CASA with microfluidics to study changes in sperm motility in a time-resolved fashion (for more details, see Supplementary Fig. 7). Mixing of control sperm from donors with progesterone evoked a rapid, transient rise in hyperactivation: Within 15 seconds, the fraction of hyperactive sperm increased by 9.4 ± 5.3% (n = 5) and then declined again within 120 seconds to basal levels (Fig. 5C). This time course indeed resembles that of progesterone-induced Ca^2+^ responses (Fig. 1C). In *CATSPER2^-/-^* sperm, the progesterone-evoked hyperactivation was abolished (Fig. 5C), demonstrating that functional CatSper is required.

**Figure 5:**
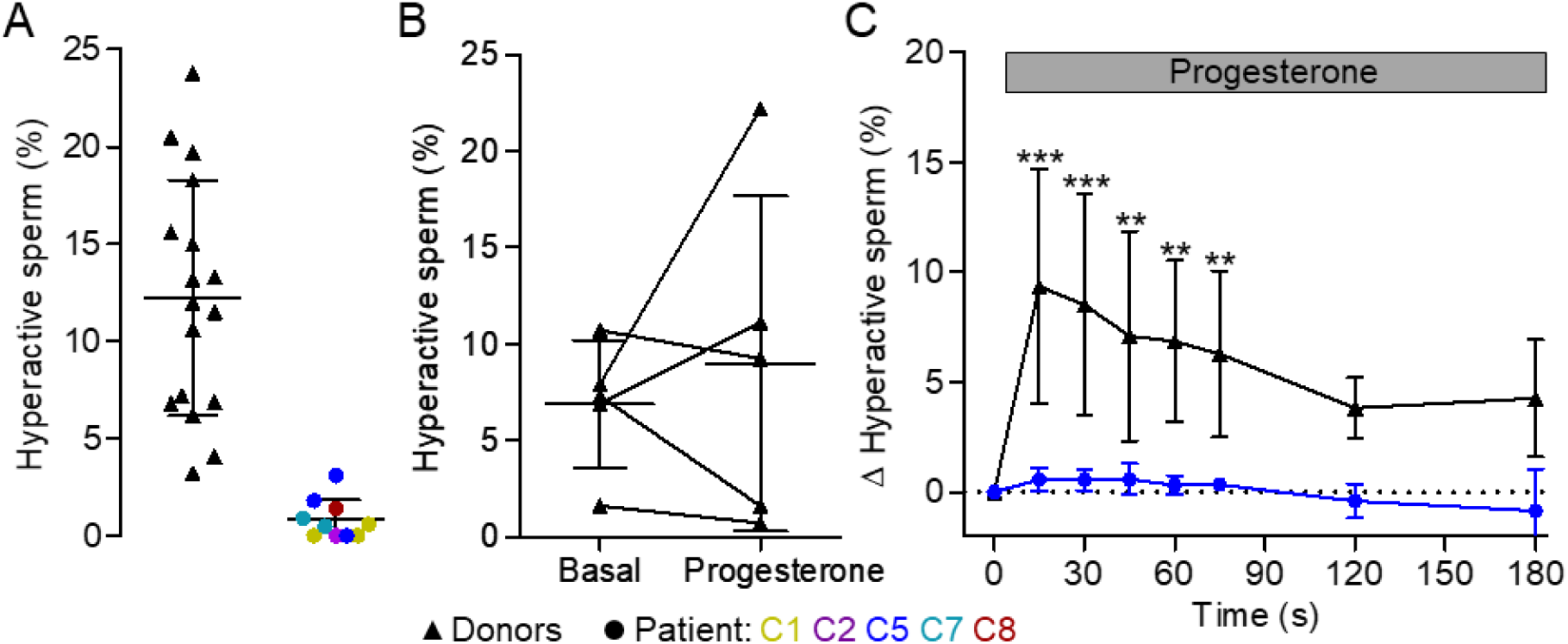
Capacitation- and progesterone-induced hyperactivation of control and *CATSPER2^-/-^* sperm. **(A)** Scatter plot (mean ± SD) of the fraction of hyperactivated control (black triangles, n = 17) and *CATSPER2^-/-^* sperm (color-coded circles, n = 10 experiments with sperm form patients C1, C2, C5, C7, and C8) upon incubation under capacitating conditions. **(B)** Paired plots of the fraction of hyperactivated control sperm from donors before (basal) and after treatment (5 min) with progesterone (5 µM) determined by standard CASA. **(C)** Change of the fraction (mean ± SD) of hyperactivated control sperm from donors (black, n = 5) and from a *CATSPER2^-/-^* patient (blue, n = 3, i.e., three independent experiments with sperm form patient C5) evoked by mixing with progesterone (gray bar), corrected for the fraction of hyperactivated sperm determined after mixing with HTF^++^ alone (set to 0 at t = 0 s), and determined by a custom kinetic CASA technique. Experiments were performed with sperm incubated under capacitating conditions. **P < 0.01, ***P < 0.001, ANOVA with Dunnett’s multiple comparison test versus the respective controls (t = 0 s).

To scrutinize this finding by an independent technique, we studied the action of progesterone on the flagellar beat of individual capacitated head-tethered sperm. To this end, we bathed the sperm in HTF^++^ fortified with caged progesterone (Kilic *et al*., 2009; Rennhack *et al*., 2018) and analyzed their motility and flagellar beat before and after uncaging progesterone with a brief UV flash. In general, for head-tethered sperm, the flagellar beat and its inherent asymmetry results in an oscillatory rotational motion around the point of attachment (Fig. 6A, B) (Saggiorato *et al*., 2017; Schiffer *et al*., 2014). We determined the rotation velocity (Fig. 6A, B, upper panels) as well as the frequency and amplitude of the oscillation (Fig. 6A, B, lower panels) in control and *CATSPER2^-/-^* sperm before and after uncaging progesterone (see also Supplementary Movie 3 and 4). Before uncaging progesterone, the *CATSPER2^-/-^* sperm featured a slightly, but statistically insignificant, enhanced beat frequency and reduced beat amplitude compared to control sperm, whereas the rotation velocity was similar (Fig. 6B, Supplementary Fig. 8). In control sperm, uncaging progesterone decreased the beat frequency by 3.1 ± 1.5 Hz (n = 9), whereas the beat amplitude and rotation velocity increased by 4.9 ± 7.4° and 95 ± 69°·s^-1^ (n = 9), respectively, suggesting a switch to hyperactive motility (Fig. 6B, Supplementary Fig. 8, Supplementary Movie 3). In *CATSPER2^-/-^*sperm, progesterone did not affect these motility parameters (Fig. 6B, Supplementary Fig. 8, Supplementary Movie 4).

**Figure 6:**
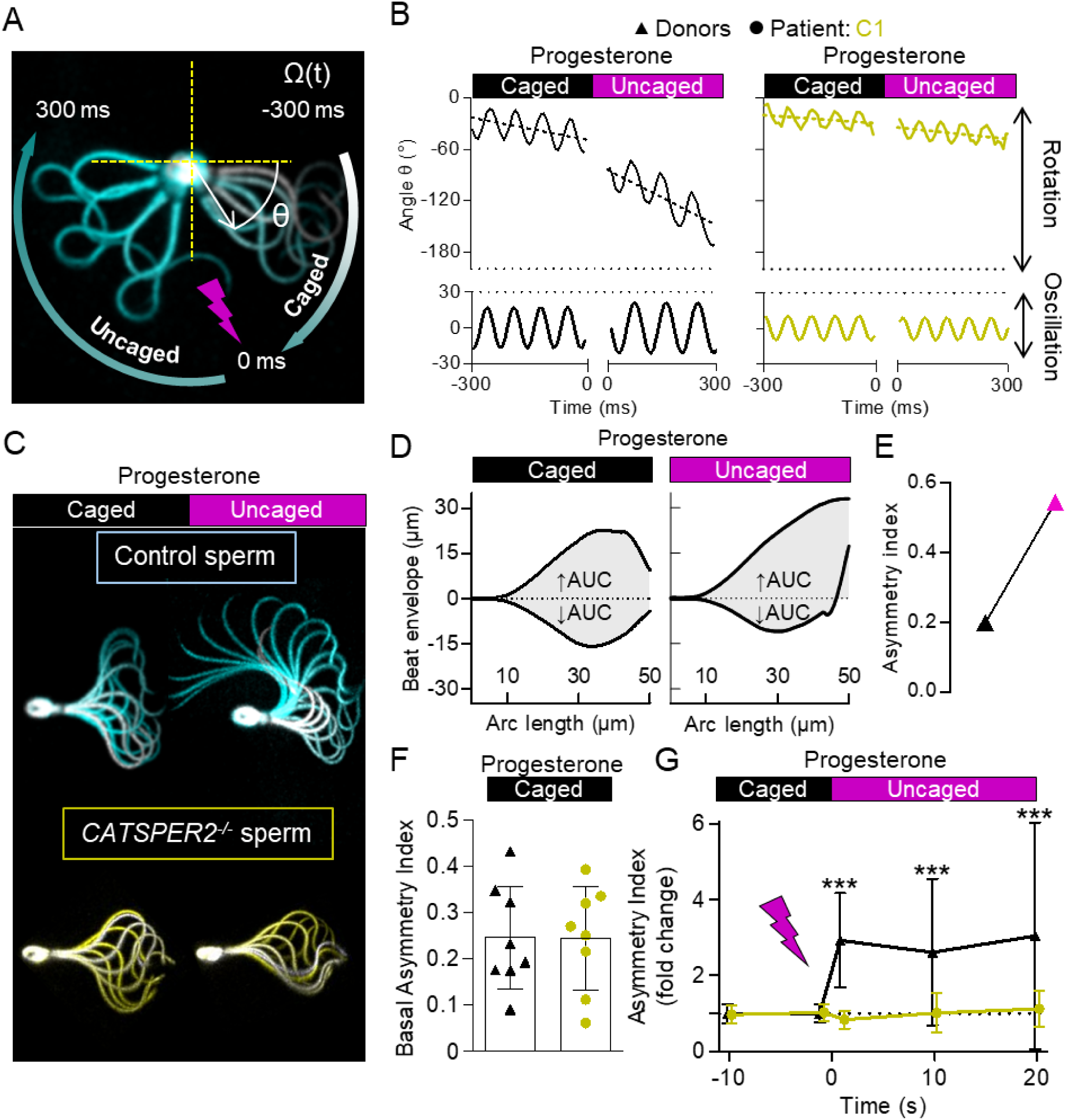
Progesterone-induced changes in flagellar beat of control and *CATSPER2^-/-^* sperm. **(A)** Representative time-lapse overlays of three beat cycles before and after uncaging progesterone (2 µM) of a pivoting head-tethered capacitated control sperm from a donor. **(B)** Change of angle θ over time (solid line, upper panels) with the corresponding slope (dotted line) and fitted sine wave of the oscillation (lower panels) of control (black, left panel) and *CATSPER2^-/-^* (gold, right panel) sperm from patient C1 before and after uncaging progesterone. The corresponding frequency and amplitude of the flagellar beat were derived from the fitted sine wave and the rotation velocity, Ω(°·s^-1^) was derived by the slope of the change of θ over time. **(C)** Representative overlays of a single beat cycle of a control (cyan) and a *CATSPER2^-/-^* (gold) sperm before and after the uncaging of progesterone (2 µM). **(D)** The area under the curve outlined by the beat envelope above (↑AUC) and below (↓AUC) the head-midpiece axis of a control sperm before (left panel) and after (right panel) the photo-release of caged progesterone used to derive an asymmetry index (for more details, see material and methods). **(E)** Paired plot depicting the corresponding asymmetry index of the flagellar beat in (D). **(F)** The asymmetry index (mean ± SD) of the basal flagellar beat of control (black, n = 8, two donors) and *CATSPER2^-/-^*(gold, n = 8, patient C1) sperm prior to progesterone exposure. **(G)** The change in the asymmetry index (mean ± SD) relative to the asymmetry index immediately prior (set to 0 at the mean of t = -10 s and 0 s) to the photo-release of progesterone of control (black triangles) and *CATSPER2^-/-^* (gold circles) sperm described in (F). ***P < 0.001, ANOVA with Dunnett’s multiple comparison test versus the respective controls (t = 0 s).

To determine whether progesterone affects also the asymmetry of the flagellar beat, we superimposed images of individual head-tethered sperm recorded during one beat cycle and analyzed these quasi stop-motion images (Fig. 6C). Before uncaging, both control (Fig. 6C, cyan) and *CATSPER2^-/-^* (Fig. 6C, gold) sperm beat largely symmetric. In control, but not in *CATSPER^-/-^* sperm, uncaging progesterone induced an increase in beat amplitude and a highly asymmetrical, whip-like beating pattern – hallmarks of hyperactive motility (Fig. 6C). The changes in beat asymmetry were quantified by determining an ‘asymmetry index’ (Fig. 6D, E); asymmetry-index values of 0 and 1 indicate perfect beat symmetry and maximal asymmetry, respectively. Prior to uncaging progesterone, the asymmetry index was similarly low in control and *CATSPER2^-/-^* sperm (0.25 ± 0.11 versus 0.25 ± 0.11, n = 8) (Fig. 6F). Progesterone increased the asymmetry index in control sperm by about ∼2.5-fold (Fig. 6D, E, G), whereas in *CATSPER2^-/-^*sperm, if at all, it slightly decreased (Fig. 6G). Altogether, the motility and flagellar-beat analyses show that loss of CatSper function abolishes both spontaneous and progesterone-induced hyperactivation.

We also studied the migration of control and *CATSPER2^-/-^*sperm into viscous medium using a modified Kremer’s sperm-mucus penetration test. An open glass capillary containing HTF^++^ fortified with methylcellulose was partially submersed in a suspension of sperm incubated under capacitating conditions in HTF^++^ (Fig. 7A). In the absence of progesterone, the numbers of control and *CATSPER2^-/-^* sperm penetrating the viscous medium were similar (Fig. 7B), demonstrating that penetration into viscous medium *per se* does not require CatSper (Luo *et al*., 2019; Williams *et al*., 2015). In the presence of progesterone, the number of control sperm was enhanced by 1.7- fold (Fig. 7B, C), confirming that progesterone facilitates the migration into viscous medium (Brown *et al*., 2017; Luo *et al*., 2019; Rahban *et al*., 2021; Rehfeld *et al*., 2020; Williams *et al*., 2015). This action of progesterone was abolished in *CATSPER2^-/-^* sperm (Fig. 7B, C), demonstrating that it requires functional CatSper (Luo *et al*., 2019; Williams *et al*., 2015).

**Figure 7:**
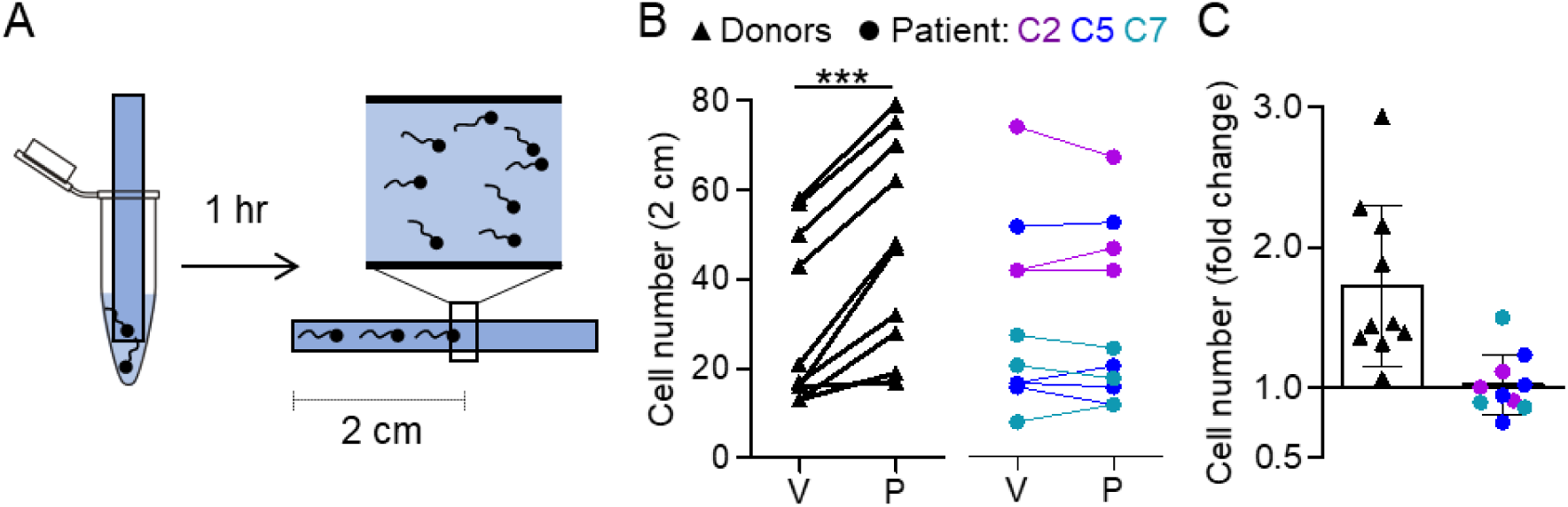
Viscous media penetration of control and *CATSPER2^-/-^* sperm. **(A)** Experimental layout of the viscous media penetration test. Glass capillaries are filled with a 1% w/v methyl cellulose solution in HTF^++^ fortified with either DMSO (vehicle) or progesterone (3 µM) and then placed in tubes containing capacitated sperm suspended in HTF^++^ fortified correspondingly with DMSO or progesterone. After one hour, the number of sperm reaching the 2-cm mark in each capillary were counted. **(B)** Paired plots of the number of control (black triangles, n = 10) and *CATSPER2^-/-^* sperm (color-coded circles, n = 10 from four independent experiments with sperm from patients C2, C5, C7) at a penetration distance of 2 cm in a modified Kremer’s sperm-mucus penetration using a capillary containing the vehicle (V) or progesterone (P; 3 µM). **(C)** Fold change in the number of control (black triangles) and *CATSPER2^-/-^* sperm (color-coded circles) at 2 cm in the presence of progesterone, relative to the vehicle (set to 1); ***P < 0.001, paired t-test.

To fertilize the egg, sperm must also undergo acrosomal exocytosis (Hirohashi & Yanagimachi, 2018). We studied acrosomal exocytosis using FITC-labelled pisum sativum agglutinin (FITC-PSA) as a marker (Fig. 8A). Spontaneous and ionomycin-induced acrosomal exocytosis was similar in control and *CATSPER2^-/-^* sperm (Fig. 8B), confirming that it is not per se affected by loss of CatSper function (Luo *et al*., 2019); see however (Wang *et al*., 2021b). To elucidate the role of Ca^2+^ influx via CatSper, we studied the action of progesterone, which in most (Harper *et al*., 2006; Krausz *et al*., 1996; Luo *et al*., 2019; Rahban *et al*., 2021; Rehfeld *et al*., 2020; Rennhack *et al*., 2018; Schaefer *et al*., 1998; Tamburrino *et al*., 2014) but not all (Prajapati *et al*., 2022; Uhler *et al*., 1992) previous studies sufficed as a physiological stimulus for acrosomal exocytosis and was abolished in sperm lacking functional CatSper (Luo *et al*., 2019). Under the experimental conditions used here, the fraction of acrosome reacted control sperm was not significantly enhanced by progesterone (Fig. 8B), which precluded scrutinizing the role of Ca^2+^-influx via CatSper for acrosomal exocytosis.

**Figure 8:**
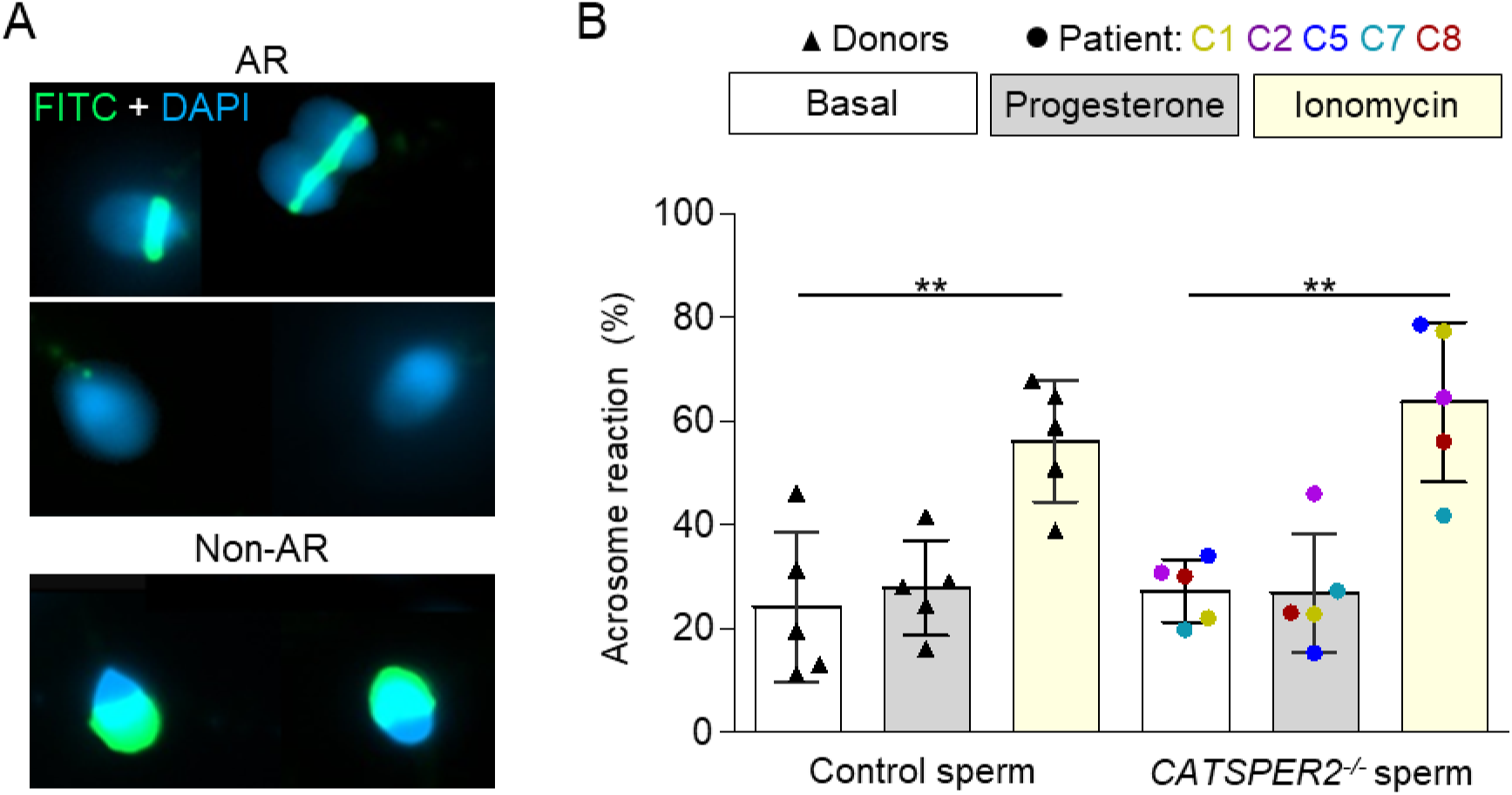
Acrosome reaction of control and *CATSPER2^-/-^* sperm. **(A)** Representative fluorescence images of capacitated control sperm from donors stained with DAPI (blue) and FITC-labelled peanut agglutinin (PNA) lectin that were considered non-acrosome reacted (non-AR) and acrosome reacted (AR). **(B)** Scatter plot of the fraction (mean ± SD) of acrosome-reacted control (black triangles, n = 5) and *CATSPER2^-/-^* sperm (color coded, n = 5) after incubation in HTF^++^ (basal; white), progesterone (5 µM; gray), or ionomycin (5 µM; yellow). Experiments were performed with sperm incubated under capacitating conditions. **P < 0.01, ANOVA with Dunnett’s multiple comparison test versus the respective basal value for control and CATSPER2*^-/-^* sperm.

## Discussion

More than two decades ago, CatSper and its essential role in sperm function and male fertility in mice were discovered (Ren *et al*., 2001). Since then, the role of CatSper in spermatogenesis, sperm function, and male fertility in humans has remained ill-defined and controversial (Brown *et al*., 2019; Wang *et al*., 2021a). To clarify these long-standing issues, we developed the CatSper-Activity-Test, screened almost 2,300 subjects, identified a group of men with defective CatSper function, assessed their reproductive phenotype, and investigated the behavior of CatSper-deficient human sperm. This shows that CatSper is not required for sperm production in men. In fact, defective CatSper function is undetectable by state-of-the-art fertility workup based on standard semen and/or computer-based sperm analysis, but causes failure of medically assisted reproduction via OI, IUI and IVF (Luo *et al*., 2019; Wang *et al*., 2021b; Williams *et al*., 2015) – techniques of primary choice for couples presenting with unexplained infertility.

The infertility and need for ICSI can readily be explained by the failure of CatSper-deficient human sperm to hyperactivate spontaneously and upon hormone stimulation and, thus, to penetrate the zona pellucida on their own. Yet, loss of CatSper function also affects migration in viscous medium *in vitro*, which could prevent the sperm to even reach the site of fertilization *in vivo*. To this end, human sperm might also be instructed by long- and short-range cues to locate the oocyte by rheotaxis, thermotaxis, chemotaxis, or a combination thereof (Alvarez *et al*., 2014)). CatSper is not required for rheotaxis (Schiffer *et al*., 2020), but the role of CatSper in human sperm chemotaxis and thermotaxis remains to be elucidated. Thus, studies mimicking *in vitro* the complex chemical, topographical, and hydrodynamic landscapes of the female genital tract (Kirkman-Brown & Smith, 2011; Suarez, 2008; Suarez & Pacey, 2006) are required to gain further insights into the role of CatSper for sperm migration and navigation *in vivo*. Future studies are also required to understand the role of Ca^2+^ influx via CatSper for acrosomal exocytosis using, e.g., physiological combinations of ligands and/or intracellular alkalization, which is more efficacious than progesterone alone in stimulating Ca^2+^ influx via CatSper (Brenker *et al*., 2018a; Prajapati *et al*., 2022) and acrosomal exocytosis (Prajapati *et al*., 2022; Schaefer *et al*., 1998). Finally, whether impaired CatSper function like in patient C9 also affects sperm function and male fertility remains to be determined. To this end, it is required to identify more patients with impaired rather than loss of CatSper function, unravel if they can naturally conceive, and perform an in-depth analysis of the phenotype of their sperm. This might reveal the critical level of CatSper activity required for natural conception and IUI and/or IVF.

We further show that homozygous deletion of *CATSPER2* is the main cause affecting CatSper function. Only in half of the *CATSPER2^-/-^*patients who we identified, *STRC* was also deleted, corresponding with the sensorineural hearing loss of affected patients. Our findings match with population genetics: Among the (likely) pathogenic variants identified in genes encoding CatSper subunits, deletion of *CATSPER2* is by far the most common one with an allele frequency of about 1% each for deletion of *CATSPER2* or *CATSPER2* + *STRC* (gnomAD SVs 2.1). The high deletion frequency is most likely due to the architecture of the particular locus: The *PPIP5K1, CKMT1, STRC*, and *CATSPER2* genes share > 98% sequence homology (Avidan *et al*., 2003) with their, except for *CKMT1*, pseudogenic counterparts created by segmental duplication (Fig. 2). This renders the genes prone to non-allelic homologous recombination (NAHR), resulting in sequence deletions, duplications, or inversions (Liu *et al*., 2012; Zhang *et al*., 2007). Of note, sperm from a man heterozygous for the deletion of *CATSPER2* (*CATSPER2*^+/-^) featured CatSper-mediated membrane currents as well as basal motility parameters similar to that of control sperm from donors (Supplementary Fig. 9), supporting fully penetrant autosomal-recessive inheritance of *CATSPER2*-related infertility. The c.2394-2399del variant in *CATSPERE* of patient C9 has already been associated with CatSper-related male infertility (Brown *et al*., 2018; Williams *et al*., 2015) and features an allele frequency of 0.01%. The c.536G>A variant in *CATSPERE* as well as those previously described for *CATSPER1* and *CATSPER3* are much less frequent or even novel.

Furthermore, from the allele frequencies in the general population, we can deduce that the prevalence for defective CatSper function due to homozygous deletion of *CATSPER2* is ∼0.01%, i.e., 1:10,000 men should be affected. In the group of men enrolled in our study, however, the prevalence was orders of magnitudes higher (Supplementary Fig. 4). This enrichment is to be expected, considering that we selected for men with fertility disorders. Nevertheless, even among the 500 men that presented for other reasons, we identified by chance two with defective CatSper function. Similarly, because of the frequent concomitant deletion of *STRC*, the prevalence for homozygous deletion of *CATSPER2* is three orders of magnitude higher in men with mild-to-moderate hearing loss than predicted for the general population (Han *et al*., 2021; Yokota *et al*., 2019).

Because CatSper-related male infertility is undetectable by standard semen analysis, affected couples experience failed OI, IUI, and IVF treatments (Table 1). This is highly frustrating, time-consuming, expensive, and, most importantly, involves repeated treatment of the female. Thus, early diagnosis of CatSper-related male-factor infertility enables an evidence-based selection of the technique for medically assisted reproduction, shortening the time towards reproductive success for affected couples, reducing both psychological burdens and expenses, and minimizing the medical risk for the female. Given its simplicity and benefit, we envision the CatSper-Activity-Test be performed along with standard semen analysis to identify patients with defective CatSper function. Importantly, its readout reveals defective CatSper function irrespective of the underlying genetic or non-genetic pathomechanism. In fact, such transfer of basic knowledge into the clinics, undoubtedly improving both diagnostic and care of patients, has been lacking in the field of male infertility. Our study might, thus, serve as a blueprint for future studies unraveling the mechanisms underlying seemingly unexplained infertility and developing novel diagnostic tools.

## Supporting information

Representative video of the ejaculate of a CATSPER2-/- patient

Video of an IVF attempt with sperm from the CATSPER2-/- patient C5

Video of head-tethered control sperm before and after uncaging progesterone

Video of head-tethered CATSPER2-/- sperm before and after uncaging progesterone

## Data Availability

All data produced in the present study are available upon reasonable request to the authors

## Acknowledgements

We thank all present and former members of the Andrological Laboratory of our center as well as the donors and patients for supporting this study. B.R., D.W., S.K., S.S., F.T., C.B. and T.S. are supported by the Deutsche Forschungsgemeinschaft (DFG, German Research Foundation) - project numbers 329621271 (CRU326; B.R., S.K., S.S., F.T., C.B., T.S,), 420653497 (CRC1450; B.R.), 432325352 (SFB1454; D.W.), 450149205 (TRR333/1; D.W.), and 390873048 (EXC2151; D.W.). D.W. is also supported by the Else-Kröner-Fresenius Foundation (2021.EKFSE.53) and intramural funding from the University of Bonn, T.S. by the Interdisciplinary Center for Clinical Research Münster (IZKF; Str/014/21) and the Europäischer Fonds für regionale Entwicklung (EFRE; 0400305), C.B. by the “Innovative Medical Research” of the University of Münster Medical School (IMF; BR 121507), and S.Y/C.S by the EXIST program of the Federal Ministry of Economic Affairs and Climate Action and European (Europäischer Sozialfonds; 03EFQNW261).

## Author contributions

C.Br., C.S., and T.S. conceived the project. S.Y., C.S., C.Br., F.T., and T.S designed the research and interpreted the results. S.Y., C.S., A.W., J. P., A.R., M.K., C. Bi., L.He., B.S., A.R., T.W., L.Ha., D.D., C.Br., and T.S. performed experiments and analyzed the data. V.N., T.P., C.K., J. N.H., D.W., B.R., H.M.B, S.S. and S.K. contributed to the design of research and/or analysis and interpretation of results. C.K., S.K. and F.T. recruited the men enrolled in the study. C.K., V.N., T.P., A.R., H.M.B., S.K., and F.T. performed the genetic and clinical workup and treatment of patients/couples. S.Y. and T.S. wrote the manuscript. C.S., C.Br., and F.T. contributed to the writing of the manuscript. All authors revised the manuscript critically for important intellectual content and approved the manuscript.

## References

Agarwal, A., Mulgund, A., Hamada, A., & Chyatte, M. R. (2015). A unique view on male infertility around the globe. Reproductive Biology and Endocrinology, 13(1), 1–9. https://doi.org/10.1186/s12958-015-0032-1

Alvarez, L., Friedrich, B. M., Gompper, G., & Kaupp, U. B. (2014). The computational sperm cell. Trends in Cell Biology, 24(3), 198–207. https://doi.org/10.1016/j.tcb.2013.10.004

Avenarius, M. R., Hildebrand, M. S., Zhang, Y., Meyer, N. C., Smith, L. L. H., Kahrizi, K., Najmabadi, H., & Smith, R. J. H. (2009). Human Male Infertility Caused by Mutations in the CATSPER1 Channel Protein. American Journal of Human Genetics, 84(4), 505–510. https://doi.org/10.1016/j.ajhg.2009.03.004

Avidan, N., Tamary, H., Dgany, O., Cattan, D., Pariente, A., Thulliez, M., Borot, N., Moati, L., Barthelme, A., Shalmon, L., Krasnov, T., Ben-Asher, E., Olender, T., Khen, M., Yaniv, I., Zaizov, R., Shalev, H., Delaunay, J., Fellous, M., Lancet, D., & Beckmann, J. S. (2003). CATSPER2, a human autosomal nonsyndromic male infertility gene. European Journal of Human Genetics, 11(7), 497–502. https://doi.org/10.1038/sj.ejhg.5200991

Baldi, E., Luconi, M., Muratori, M., Marchiani, S., Tamburrino, L., & Forti, G. (2009). Nongenomic activation of spermatozoa by steroid hormones: Facts and fictions. Molecular and Cellular Endocrinology, 308(1–2), 39–46. https://doi.org/10.1016/j.mce.2009.02.006

Barratt, C. L. R., Mansell, S., Beaton, C., Tardif, S., & Oxenham, S. K. (2011). Diagnostic tools in male infertility-the question of sperm dysfunction. Asian Journal of Andrology, 13(1), 53–58. https://doi.org/10.1038/aja.2010.63

Brenker, C., Rehfeld, A., Schiffer, C., Kierzek, M., Kaupp, U. B., Skakkebæk, N. E., & Strünker, T. (2018a). Synergistic activation of CatSper Ca2+ channels in human sperm by oviductal ligands and endocrine disrupting chemicals. Human Reproduction, 33(10), 1915–1923. https://doi.org/10.1093/humrep/dey275

Brenker, C., Schiffer, C., Wagner, I. V., Tüttelmann, F., Röpke, A., Rennhack, A., Kaupp, U. B., & Strünker, T. (2018b). Action of steroids and plant triterpenoids on CatSper Ca ^2+^ channels in human sperm. Proceedings of the National Academy of Sciences, 201717929. https://doi.org/10.1073/pnas.1717929115

Brown, S. G., Costello, S., Kelly, M. C., Ramalingam, M., Drew, E., Publicover, S. J., Barratt, C. L. R., & Da Silva, S. M. (2017). Complex CatSper-dependent and independent [Ca2+] i signalling in human spermatozoa induced by follicular fluid. Human Reproduction, 32(10), 1995–2006. https://doi.org/10.1093/humrep/dex269

Brown, S. G., Miller, M. R., Lishko, P. V., Lester, D. H., Publicover, S. J., Barratt, C. L. R., & Da Silva, S. M. (2018). Homozygous in-frame deletion in CATSPERE in a man producing spermatozoa with loss of CatSper function and compromised fertilizing capacity. Human Reproduction, 33(10), 1812–1816. https://doi.org/10.1093/humrep/dey278

Brown, S. G., Publicover, S. J., Barratt, C. L. R., & Martins da Silva, S. J. (2019). Human sperm ion channel (dys)function: Implications for fertilization. Human Reproduction Update, 25(6), 758–776. https://doi.org/10.1093/humupd/dmz032

Carlson, A. E., Quill, T. A., Westenbroek, R. E., Schuh, S. M., Hille, B., & Babcock, D. F. (2005). Identical phenotypes of CatSper1 and CatSper2 null sperm. Journal of Biological Chemistry, 280(37), 32238–32244. https://doi.org/10.1074/jbc.M501430200

Carlson, A. E., Westenbroek, R. E., Quill, T., Ren, D., Clapham, D. E., Hille, B., Garbers, D. L., & Babcock, D. F. (2003). CatSper1 required for evoked Ca2+ entry and control of flagellar function in sperm. Proceedings of the National Academy of Sciences, 100(25), 14864– 14868. https://doi.org/10.1073/pnas.2536658100

Chung, J.-J., Miki, K., Kim, D., Shim, S. H., Shi, H. F., Hwang, J. Y., Cai, X., Iseri, Y., Zhuang, X., & Clapham, D. E. (2017). Catsperς regulates the structural continuity of sperm ca2+ signaling domains and is required for normal fertility. ELife, 6, 1–25. https://doi.org/10.7554/eLife.23082

Chung, J.-J., Navarro, B., Krapivinsky, G., Krapivinsky, L., & Clapham, D. E. (2011). A novel gene required for male fertility and functional CATSPER channel formation in spermatozoa. Nature Communications, 2(1), 153. https://doi.org/10.1038/ncomms1153

Chung, J.-J., Shim, S. H., Everley, R. A., Gygi, S. P., Zhuang, X., & Clapham, D. E. (2014). Structurally distinct Ca2+ signaling domains of sperm flagella orchestrate tyrosine phosphorylation and motility. Cell, 157(4), 808–822. https://doi.org/10.1016/j.cell.2014.02.056

Giaretta, E., Munerato, M., Yeste, M., Galeati, G., Spinaci, M., Tamanini, C., Mari, G., & Bucci, D. (2017). Implementing an open-access CASA software for the assessment of stallion sperm motility: Relationship with other sperm quality parameters. Animal Reproduction Science, 176, 11–19. https://doi.org/10.1016/j.anireprosci.2016.11.003

Han, S., Zhang, D., Guo, Y., Fu, Z., & Guan, G. (2021). Prevalence and Characteristics of STRC Gene Mutations (DFNB16): A Systematic Review and Meta-Analysis. Frontiers in Genetics, 12(September). https://doi.org/10.3389/fgene.2021.707845

Hansen, J., Rassmann, S., Jikeli, J., & Wachten, D. (2018). SpermQ–A Simple Analysis Software to Comprehensively Study Flagellar Beating and Sperm Steering. Cells, 8(1), 10. https://doi.org/10.3390/cells8010010

Harper, C. V., Barratt, C. L. R., Publicover, S. J., & Kirkman-Brown, J. C. (2006). Kinetics of the Progesterone-Induced Acrosome Reaction and Its Relation to Intracellular Calcium Responses in Individual Human Spermatozoa1. Biology of Reproduction, 75(6), 933–939. https://doi.org/10.1095/biolreprod.106.054627

Hildebrand, M. S., Avenarius, M. R., Fellous, M., Zhang, Y., Meyer, N. C., Auer, J., Serres, C., Kahrizi, K., Najmabadi, H., Beckmann, J. S., & Smith, R. J. H. (2010). Genetic male infertility and mutation of CATSPER ion channels. European Journal of Human Genetics, 18(11), 1178–1184. https://doi.org/10.1038/ejhg.2010.108

Hirohashi, N., & Yanagimachi, R. (2018). Sperm acrosome reaction: Its site and role in fertilization. Biology of Reproduction, 99(1), 127–133. https://doi.org/10.1093/biolre/ioy045

Ho, K., Wolff, C. A., & Suarez, S. S. (2009). CatSper-null mutant spermatozoa are unable to ascend beyond the oviductal reservoir. *Reproduction*, Fertility and Development, 21(2), 345–350. https://doi.org/10.1071/RD08183

Hwang, J. Y., Mannowetz, N., Zhang, Y., Everley, R. A., Gygi, S. P., Bewersdorf, J., Lishko, P. V., & Chung, J.-J. (2019). Dual Sensing of Physiologic pH and Calcium by EFCAB9 Regulates Sperm Motility. Cell, 1–15. https://doi.org/10.1016/j.cell.2019.03.047

Hwang, J. Y., Wang, H., Lu, Y., Ikawa, M., & Chung, J.-J. (2022). C2cd6-encoded CatSperτ targets sperm calcium channel to Ca2+ signaling domains in the flagellar membrane. Cell Reports, 110226. https://doi.org/10.1016/j.celrep.2021.110226

Jeschke, J. K., Biagioni, C., Schierling, T., Wagner, I. V., Börgel, F., Schepmann, D., Schüring, A., Kulle, A. E., Holterhus, P. M., von Wolff, M., Wünsch, B., Nordhoff, V., Strünker, T., & Brenker, C. (2021). The Action of Reproductive Fluids and Contained Steroids, Prostaglandins, and Zn2+ on CatSper Ca2+ Channels in Human Sperm. Frontiers in Cell and Developmental Biology, 9(July), 1–16. https://doi.org/10.3389/fcell.2021.699554

Jin, J., Jin, N., Zheng, H., Ro, S., Tafolla, D., Sanders, K. M., & Yan, W. (2007). Catsper3 and Catsper4 are essential for sperm hyperactivated motility and male fertility in the mouse. Biology of Reproduction, 77(1), 37–44. https://doi.org/10.1095/biolreprod.107.060186

Kilic, F., Kashikar, N. D., Schmidt, R., Alvarez, L., Dai, L., Weyand, I., Wiesner, B., Goodwin, N., & Hagen, V. (2009). Caged Progesterone : A New Tool for Studying Rapid Nongenomic Actions of Progesterone. JACS, 12, 4027–4030. https://doi.org/10.1021/ja808334f

Kirkman-Brown, J. C., & Smith, D. J. (2011). Sperm motility: Is viscosity fundamental to progress? Molecular Human Reproduction, 17(8), 539–544. https://doi.org/10.1093/molehr/gar043

Krausz, C., Bonaccorsi, L., Maggio, P., Luconi, M., Criscuoli, L., Fuzzi, B., Pellegrini, S., Forti, G., & Baldi, E. (1996). Two functional assays of sperm responsiveness to progesterone and their predictive values in in-vitro fertilization. Human Reproduction, 11(8), 1661–1667. https://doi.org/10.1093/oxfordjournals.humrep.a019466

Lin, S., Ke, M., Zhang, Y., Yan, Z., & Wu, J. (2021). Structure of a mammalian sperm cation channel complex. Nature, 595(March). https://doi.org/10.1038/s41586-021-03742-6

Lishko, P. V., Botchkina, I. L., & Kirichok, Y. (2011). Progesterone activates the principal Ca2+ channel of human sperm. Nature, 471(7338), 387–392. https://doi.org/10.1038/nature09767

Lishko, P. V., & Kirichok, Y. (2010). The role of Hv1 and CatSper channels in sperm activation. Journal of Physiology, 588(23), 4667–4672. https://doi.org/10.1113/jphysiol.2010.194142

Liu, J., Xia, J., Cho, K. H., Clapham, D. E., & Ren, D. (2007). CatSperβ, a novel transmembrane protein in the CatSper channel complex. Journal of Biological Chemistry, 282(26), 18945– 18952. https://doi.org/10.1074/jbc.M701083200

Liu, P., Carvalho, C. M. B., Hastings, P. J., & Lupski, J. R. (2012). Mechanisms for recurrent and complex human genomic rearrangements. Current Opinion in Genetics and Development, 22(3), 211–220. https://doi.org/10.1016/j.gde.2012.02.012

Loux, S. C., Crawford, K. R., Ing, N. H., González-Fernández, L., Macías-García, B., Love, C. C., Varner, D. D., Velez, I. C., Choi, Y. H., & Hinrichs, K. (2013). CatSper and the Relationship of hyperactivated motility to intracellular calcium andpH kinetics in equine sperm. Biology of Reproduction, 89(5), 1–15. https://doi.org/10.1095/biolreprod.113.111708

Luo, T., Chen, H., Zou, Q., Wang, T., Cheng, Y., Wang, H., Wang, F., Jin, Z., Chen, Y., Weng, S., & Zeng, X. (2019). A novel copy number variation in *CATSPER2* causes idiopathic male infertility with normal semen parameters. Human Reproduction, 1–10. https://doi.org/10.1093/humrep/dey377

Luque, G. M., Xu, X., Romarowski, A., Gervasi, M. G., Orta, G., De la Vega-Beltrán, J. L., Stival, C., Gilio, N., Dalotto-Moreno, T., Krapf, D., Visconti, P. E., Krapf, D., Darszon, A., & Buffone, M. G. (2021). Cdc42 localized in the CatSper signaling complex regulates cAMP-dependent pathways in mouse sperm. FASEB Journal, 35(8), 1–21. https://doi.org/10.1096/fj.202002773RR

Mortimer, S. T., Swan, M. A., & Mortimer, D. (1998). Effect of seminal plasma on capacitation and hyperactivation in human spermatozoa. Human Reproduction, 13(8), 2139–2146. https://doi.org/10.1093/humrep/13.8.2139

Navarro, B., Kirichok, Y., Chung, J.-J., & Clapham, D. E. (2008). Ion Channels that Control Fertility in Mammalian Spermatozoa. International Journal of Developmental Biology, 52, 607–613. https://doi.org/10.1387/ijdb.072554bn.

Prajapati, P., Kane, S., McBrinn, R. C., Dean, M. S., Martins da Silva, S. J., & Brown, S. G. (2022). Elevated and Sustained Intracellular Calcium Signalling Is Necessary for Efficacious Induction of the Human Sperm Acrosome Reaction. International Journal of Molecular Sciences, 23(19). https://doi.org/10.3390/ijms231911253

Publicover, S. J., Giojalas, L. C., Teves, M. E., de Oliveira, G. S. M. M., Garcia, A. A. M., Barratt, C. L. R., & Harper, C. V. (2008). Ca2+ signalling in the control of motility and guidance in mammalian sperm. Frontiers in Bioscience : A Journal and Virtual Library, 13(April 2016), 5623–5637. https://doi.org/10.2741/3105

Qi, H., Moran, M. M., Navarro, B., Chong, J. A., Krapivinsky, G., Krapivinsky, L., Kirichok, Y., Ramsey, I. S., Quill, T. A., & Clapham, D. E. (2007). All four CatSper ion channel proteins are required for male fertility and sperm cell hyperactivated motility. Proceedings of the National Academy of Sciences, 104(4), 1219–1223. https://doi.org/10.1073/pnas.0610286104

Quill, T. A., Ren, D., Clapham, D. E., & Garbers, D. L. (2001). A voltage-gated ion channel expressed specifically in spermatozoa. Proceedings of the National Academy of Sciences, 98(22), 12527–12531. https://doi.org/10.1073/pnas.221454998

Quill, T. A., Sugden, S. A., Rossi, K. L., Doolittle, L. K., Hammer, R. E., & Garbers, D. L. (2003). Hyperactivated sperm motility driven by CatSper2 is required for fertilization. Proceedings of the National Academy of Sciences, 100(25), 14869–14874. https://doi.org/10.1073/pnas.2136654100

Rahban, R., & Nef, S. (2020). CatSper: The complex main gate of calcium entry in mammalian spermatozoa. Molecular and Cellular Endocrinology, 518(April), 110951. https://doi.org/10.1016/j.mce.2020.110951

Rahban, R., Rehfeld, A., Schiffer, C., Brenker, C., Egeberg Palme, D. L., Wang, T., Lorenz, J., Almstrup, K., Skakkebaek, N. E., Strünker, T., & Nef, S. (2021). The antidepressant Sertraline inhibits CatSper Ca2+channels in human sperm. Human Reproduction, 36(10), 2638–2648. https://doi.org/10.1093/humrep/deab190

Rehfeld, A., Andersson, A. M., & Skakkebæk, N. E. (2020). Bisphenol A Diglycidyl Ether ( BADGE ) and Bisphenol Analogs, but Not Bisphenol A ( BPA ), Activate the CatSper Ca 2 + Channel in Human Sperm. 11(May), 1–11. https://doi.org/10.3389/fendo.2020.00324

Ren, D., Navarro, B., Perez, G., Jackson, A. C., Hsu, S., Shi, Q., Tilly, J. L., & Clapham, D. E. (2001). A sperm ion channel required for sperm motility and male fertility. Nature, 413(6856), 603–609. https://doi.org/10.1038/35098027

Rennhack, A., Schiffer, C., Brenker, C., Fridman, D., Nitao, E. T., Cheng, Y. M., Tamburrino, L., Balbach, M., Stölting, G., Berger, T. K., Kierzek, M., Alvarez, L., Wachten, D., Zeng, X. H., Baldi, E., Publicover, S. J., Benjamin Kaupp, U., & Strünker, T. (2018). A novel cross-species inhibitor to study the function of CatSper Ca2+channels in sperm. British Journal of Pharmacology, 175(15), 3144–3161. https://doi.org/10.1111/bph.14355

Saggiorato, G., Alvarez, L., Jikeli, J. F., Kaupp, U. B., Gompper, G., & Elgeti, J. (2017). Human sperm steer with second harmonics of the flagellar beat. Nature Communications, 8(1). https://doi.org/10.1038/s41467-017-01462-y

Schaefer, M., Hofmann, T., Schultz, G., & Gudermann, T. (1998). A new prostaglandin E receptor mediates calcium influx and acrosome reaction in human spermatozoa. Proceedings of the National Academy of Sciences of the United States of America, 95(6), 3008–3013. https://doi.org/10.1073/pnas.95.6.3008

Schiffer, C., Müller, A., Egeberg, D. L., Alvarez, L., Brenker, C., Rehfeld, A., Frederiksen, H., Wäschle, B., Kaupp, U. B., Balbach, M., Wachten, D., Skakkebaek, N. E., Almstrup, K., & Strunker, T. (2014). Direct action of endocrine disrupting chemicals on human sperm. EMBO Reports, 13(5), 398–403.

Schiffer, C., Rieger, S., Brenker, C., Young, S., Hamzeh, H., Wachten, D., Tüttelmann, F., Röpke, A., Kaupp, U. B., Wang, T., Wagner, A., Krallmann, C., Kliesch, S., Fallnich, C., & Strünker, T. (2020). Rotational motion and rheotaxis of human sperm do not require functional CatSper channels and transmembrane Ca 2+ signaling. The EMBO Journal, 39(4), 1–15. https://doi.org/10.15252/embj.2019102363

Seifert, R., Flick, M., Bönigk, W., Alvarez, L., Trötschel, C., Poetsch, A., Müller, A., Goodwin, N., Pelzer, P., Kashikar, N. D., Kremmer, E., Jikeli, J., Timmermann, B., Kuhl, H., Fridman, D., Windler, F., Kaupp, U. B., & Strünker, T. (2015). The CatSper channel controls chemosensation in sea urchin sperm. The EMBO Journal, 34(3), 379–392. https://doi.org/10.15252/embj.201489376

Smith, J. F., Syritsyna, O., Fellous, M., Serres, C., Mannowetz, N., Kirichok, Y., & Lishko, P. V. (2013). Disruption of the principal, progesterone-activated sperm Ca2+ channel in a CatSper2-deficient infertile patient. Proceedings of the National Academy of Sciences, 110(17), 6823–6828. https://doi.org/10.1073/pnas.1216588110

Strünker, T., Goodwin, N., Brenker, C., Kashikar, N. D., Weyand, I., Seifert, R., & Kaupp, U. B. (2011). The CatSper channel mediates progesterone-induced Ca2+ influx in human sperm. Nature, 471(7338), 382–386. https://doi.org/10.1038/nature09769

Suarez, S. S. (2008). Control of hyperactivation in sperm. Human Reproduction Update, 14(6), 647–657. https://doi.org/10.1093/humupd/dmn029

Suarez, S. S., & Pacey, A. A. (2006). Sperm transport in the female reproductive tract. Human Reproduction Update, 12(1), 23–37. https://doi.org/10.1093/humupd/dmi047

Tamburrino, L., Marchiani, S., Minetti, F., Forti, G., Muratori, M., & Baldi, E. (2014). The CatSper calcium channel in human sperm: Relation with motility and involvement in progesterone-induced acrosome reaction. Human Reproduction, 29(3), 418–428. https://doi.org/10.1093/humrep/det454

Torres-Flores, V., Picazo-Juárez, G., Hernández-Rueda, Y., Darszon, A., & Gonzlez-Martínez, M. T. (2011). Sodium influx induced by external calcium chelation decreases human sperm motility. Human Reproduction, 26(10), 2626–2635. https://doi.org/10.1093/humrep/der237

Tüttelmann, F., Luetjens, C. M., & Nieschlag, E. (2006). Optimising workflow in andrology: A new electronic patient record and database. Asian Journal of Andrology, 8(2), 235–241. https://doi.org/10.1111/j.1745-7262.2006.00131.x

Tüttelmann, F., Ruckert, C., & Röpke, A. (2018). Disorders of spermatogenesis: perspectives for novel genetic diagnostics after 20 years of unchanged routine. Medizinische Genetik, 30(1), 12–20. https://doi.org/10.1007/s11825-018-0181-7

Uhler, M. L., Leung, A., Chan, S. Y. W., & Wang, C. (1992). Direct effects of progesterone and antiprogesterone on human sperm hyperactivated motility and acrosome reaction. Fertility and Sterility, 58(6), 1191–1198. https://doi.org/10.1016/S0015-0282(16)55568-X

Verpy, E., Masmoudi, S., Zwaenepoel, I., Leibovici, M., Hutchin, T. P., Del Castillo, I., Nouaille, S., Blanchard, S., Lainé, S., Popot, J. L., Moreno, F., Mueller, R. F., & Petit, C. (2001). Mutations in a new gene encoding a protein of the hair bundle cause non-syndromic deafness at the DFNB16 locus. Nature Genetics, 29(3), 345–349. https://doi.org/10.1038/ng726

Vona, B., Hofrichter, M. A. H., Neuner, C., Schröder, J., Gehrig, A., Hennermann, J. B., Kraus, F., Shehata-Dieler, W., Klopocki, E., Nanda, I., & Haaf, T. (2015). DFNB16 is a frequent cause of congenital hearing impairment: Implementation of STRC mutation analysis in routine diagnostics. Clinical Genetics, 87(1), 49–55. https://doi.org/10.1111/cge.12332

Wang, H., Liu, J., Cho, K.-H., & Ren, D. (2009). A Novel, Single, Transmembrane Protein CATSPERG Is Associated with CATSPER1 Channel Protein1. Biology of Reproduction, 81(3), 539–544. https://doi.org/10.1095/biolreprod.109.077107

Wang, H., McGoldrick, L. L., & Chung, J. J. (2021a). Sperm ion channels and transporters in male fertility and infertility. Nature Reviews Urology, 18(1), 46–66. https://doi.org/10.1038/s41585-020-00390-9

Wang, J., Tang, H., Zou, Q., Zheng, A., Li, H., Yang, S., & Xiang, J. (2021b). Patient with CATSPER3 mutations-related failure of sperm acrosome reaction with successful pregnancy outcome from intracytoplasmic sperm injection (ICSI). Molecular Genetics and Genomic Medicine, 9(2), 1–9. https://doi.org/10.1002/mgg3.1579

Williams, H. L., Mansell, S., Alasmari, W., Brown, S. G., Wilson, S. M., Sutton, K. A., Miller, M. R., Lishko, P. V, Barratt, C. L. R., Publicover, S. J., & Martins da Silva, S. (2015). Specific loss of CatSper function is sufficient to compromise fertilizing capacity of human spermatozoa. Human Reproduction, 30(12), 2737–2746. https://doi.org/10.1093/humrep/dev243

Wilson-Leedy, J. G., & Ingermann, R. L. (2006). Development of a novel CASA system based on open source software for characterization of zebrafish sperm motility parameters. Theriogenology, 67(3), 661–672. https://doi.org/10.1016/j.theriogenology.2006.10.003

Wyrwoll, M. J., Köckerling, N., Vockel, M., Dicke, A. K., Rotte, N., Pohl, E., Emich, J., Wöste, M., Ruckert, C., Wabschke, R., Seggewiss, J., Ledig, S., Tewes, A. C., Stratis, Y., Cremers, J. F., Wistuba, J., Krallmann, C., Kliesch, S., Röpke, A., Stallmeyer, B., Friedrich, C., & Tüttelmann, F. (2022). Genetic Architecture of Azoospermia—Time to Advance the Standard of Care. European Urology. https://doi.org/10.1016/j.eururo.2022.05.011

Wyrwoll, M. J., Temel, Ş. G., Nagirnaja, L., Oud, M. S., Lopes, A. M., van der Heijden, G. W., Heald, J. S., Rotte, N., Wistuba, J., Wöste, M., Ledig, S., Krenz, H., Smits, R. M., Carvalho, F., Gonçalves, J., Fietz, D., Türkgenç, B., Ergören, M. C., Çetinkaya, M., Başar, M., Kahraman, S., McEleny, K., Xavier, M. J., Turner, H., Pilatz, A., Röpke, A., Dugas, M., Kliesch, S., Neuhaus, N., GEMINI Consotrium, Aston, K. I., Conrad, D. F., Veltman, J. A., Friedrich, C., & Tüttelmann, F. (2020). Bi-allelic Mutations in M1AP Are a Frequent Cause of Meiotic Arrest and Severely Impaired Spermatogenesis Leading to Male Infertility. American Journal of Human Genetics, 107(2), 342–351. https://doi.org/10.1016/j.ajhg.2020.06.010

Yokota, Y., Moteki, H., Nishio, S. ya, Yamaguchi, T., Wakui, K., Kobayashi, Y., Ohyama, K., Miyazaki, H., Matsuoka, R., Abe, S., Kumakawa, K., Takahashi, M., Sakaguchi, H., Uehara, N., Ishino, T., Kosho, T., Fukushima, Y., & Usami, S. (2019). Frequency and clinical features of hearing loss caused by STRC deletions. Scientific Reports, 9(1), 1–9. https://doi.org/10.1038/s41598-019-40586-7

Zeng, X. H., Navarro, B., Xia, X. M., Clapham, D. E., & Lingle, C. J. (2013). Simultaneous knockout of slo3 and catsper1 abolishes all alkalization- and voltage-activated current in mouse spermatozoa. Journal of General Physiology, 142(3), 305–313. https://doi.org/10.1085/jgp.201311011

Zhang, Y., Malekpour, M., Al-Madani, N., Kahrizi, K., Zanganeh, M., Mohseni, M., Mojahedi, F., Daneshi, A., Najmabadi, H., & Smith, R. J. H. (2007). Sensorineural deafness and male infertility: A contiguous gene deletion syndrome. Journal of Medical Genetics, 44(4), 233–240. https://doi.org/10.1136/jmg.2006.045765

