## Supplementary Figures and Tables for "Unexplained infertility is frequently caused by defective CatSper function preventing sperm from penetrating the egg coat"

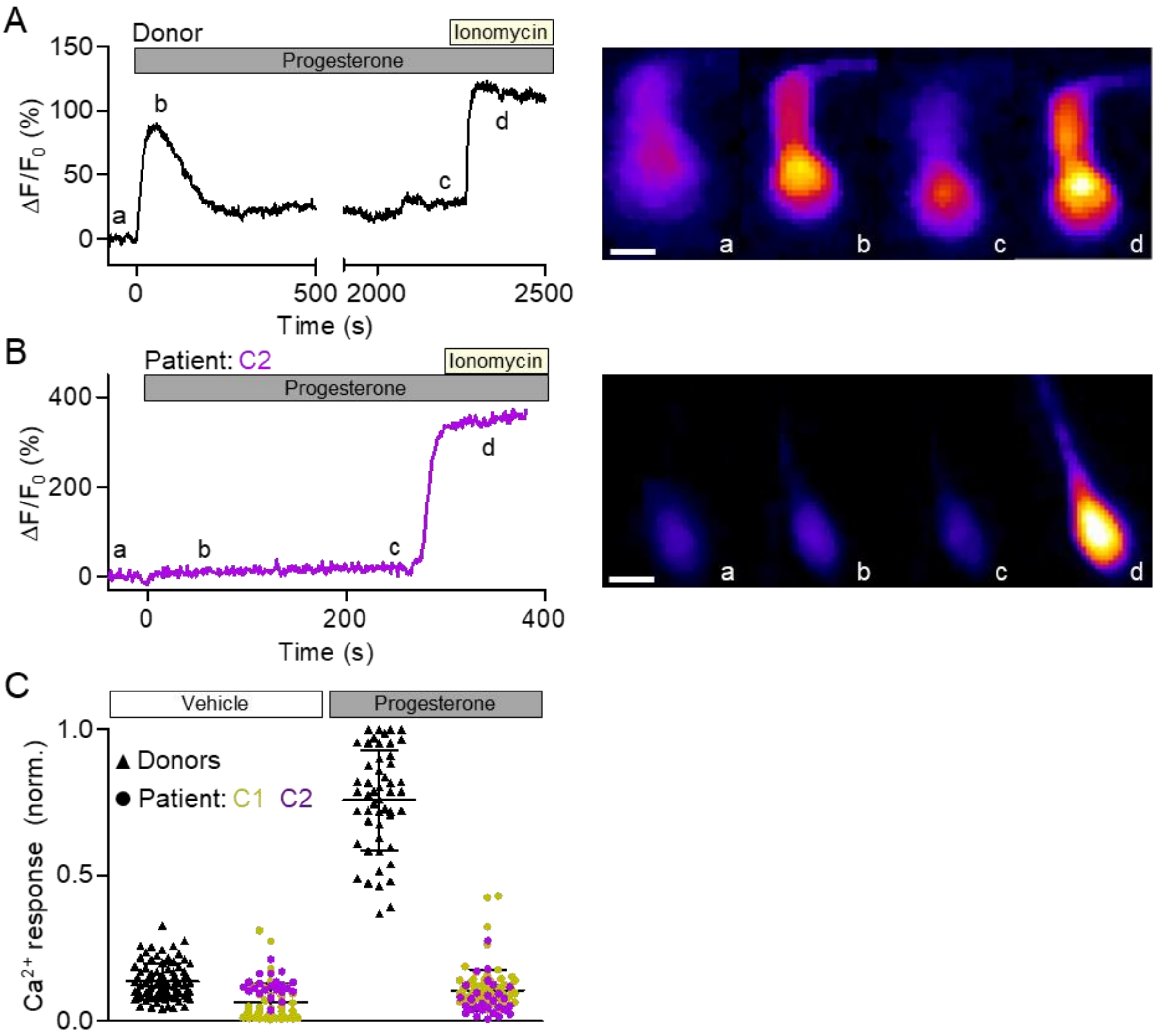


**Supplementary Figure 1: Single-cell imaging of progesterone-evoked $Ca^{2+}$ signals in control and *CATSPER2*$^{-/-}$ sperm. (A) Left:** Representative $Ca^{2+}$ response evoked by progesterone (applied at t = 0) and, subsequently, ionomycin in an immobilized sperm cell from a donor loaded with the $Ca^{2+}$ indicator Fluo-4. The signal is displayed as $\Delta F/F_0$ (%), i.e. the change in fluorescence relative to the mean basal fluorescence ($F_0$) of the ≥ 60 images before application of progesterone. **Right**: False-color coded fluorescence images taken at the indicated time points (a-d), i.e. before and at the peak of the progesterone- and ionomycin- induced $Ca^{2+}$ signal, respectively. **(B) Left:** Representative $Ca^{2+}$ response evoked by progesterone (applied at t = 0) and, subsequently, ionomycin in an immobilized *CATSPER2*$^{-/-}$ sperm cell from patient C2. **Right**: False-color coded fluorescence images taken at the indicated time points (a-d). **(C)** Scatter plot (mean ± SD) of the maximal signal amplitude evoked by application of buffer or progesterone in control (black, buffer and progesterone n = 86 and n = 51 sperm, respectively) and

*CATSPER2*$^{-/-}$ sperm (color-coded, buffer and progesterone n = 84 and n = 91 sperm, respectively) from patients C1 and C2, relative to that evoked by ionomycin (set to 1). Scale bars represent 10 µm.

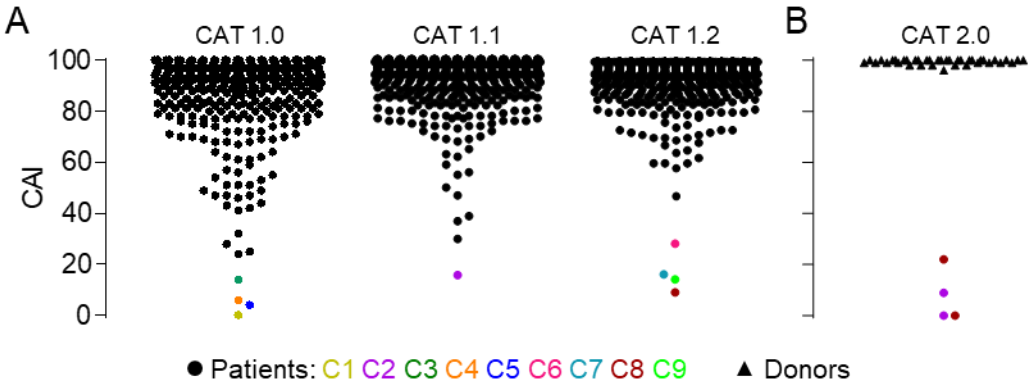


**Supplementary Figure 2: Iterative improvements of the CatSper-Activity-Test. (A)** Aggregated CatSper-Activity-Test data shown in Figure 1B separated by distinct refinement steps: CAI values from semen samples of men undergoing semen analysis determined 15 minutes (CAT 1.0, n = 370) or 30 minutes (CAT 1.1, n = 540) after dilution in Buffer A and Buffer B, or 30 minutes after dilution in Buffer B also containing EDTA (CAT 1.2, n = 1376). Patients with confirmed loss or impaired CatSper function are labeled C1-C9 and indicated with a colored-coded circle. **(B)** CAI values determined 60 minutes after dilution of semen samples from donors (black triangles, n = 38) and patients C2 and C8 (color-coded circles, n = 4) in Buffer A and Buffer B containing EDTA.

| Patient | Genotype | Patient | Genotype | Patient | Genotype |
|---|---|---|---|---|---|
| C1 | *CATSPER2*$^{-/-}$, *STRC*$^{-/-}$ | C4 | *CATSPER2*$^{-/-}$, *STRC*$^{-/-}$ | C7 | *CATSPER2*$^{-/-}$, *STRC*$^{+/-}$ |
| C2 | *CATSPER2*$^{-/-}$, *STRC*$^{-/-}$ | C5 | *CATSPER2*$^{-/-}$, *STRC*$^{+/+}$ | C8 | *CATSPER2*$^{-/-}$, *STRC*$^{+/-}$ |
| C3 | *CATSPER2*$^{-/-}$, *STRC*$^{-/-}$ | C6 | *CATSPER2*$^{-/-}$, *STRC*$^{+/-}$ | C9 | *CATSPER2*$^{+/+}$ *STRC*$^{+/+}$ |

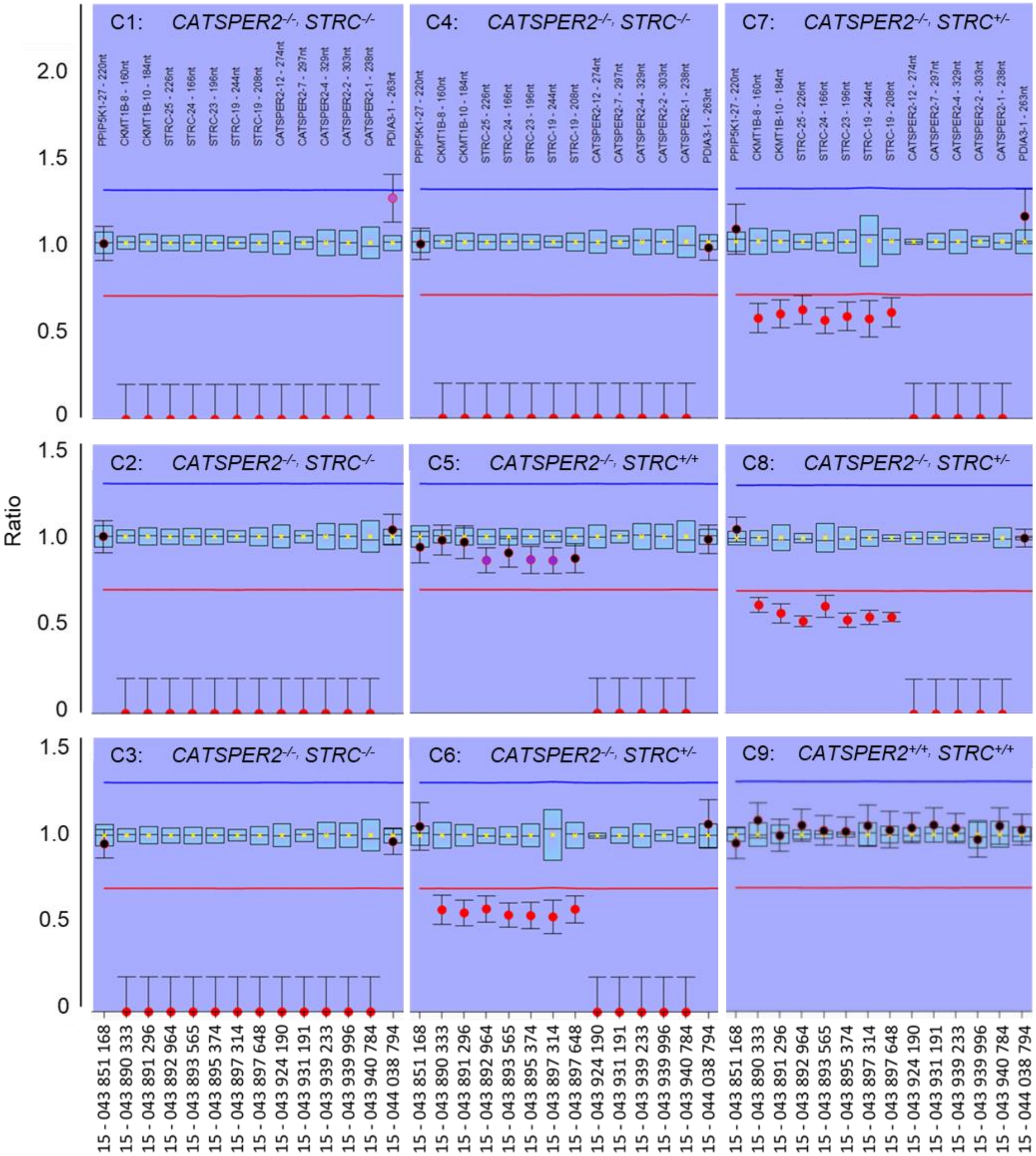


**Supplementary Figure 3: MLPA data at the chromosome 15q15.3 locus of the eight *CATSPER2*$^{-/-}$ patients.** Two flanking probes encompass reference probes for *CATSPER2*, *STRC*, and *CKMT1B*. The ratio (y-axis, range: 2.0–0) is indicative of the copy number status (normal = 0.8–1.2, heterozygous deletion = 0.4–0.7, homozygous deletion = 0). Red circles (mean ± SD) represent the patient DNA samples. Positive control DNA samples are represented as high-low box plots.

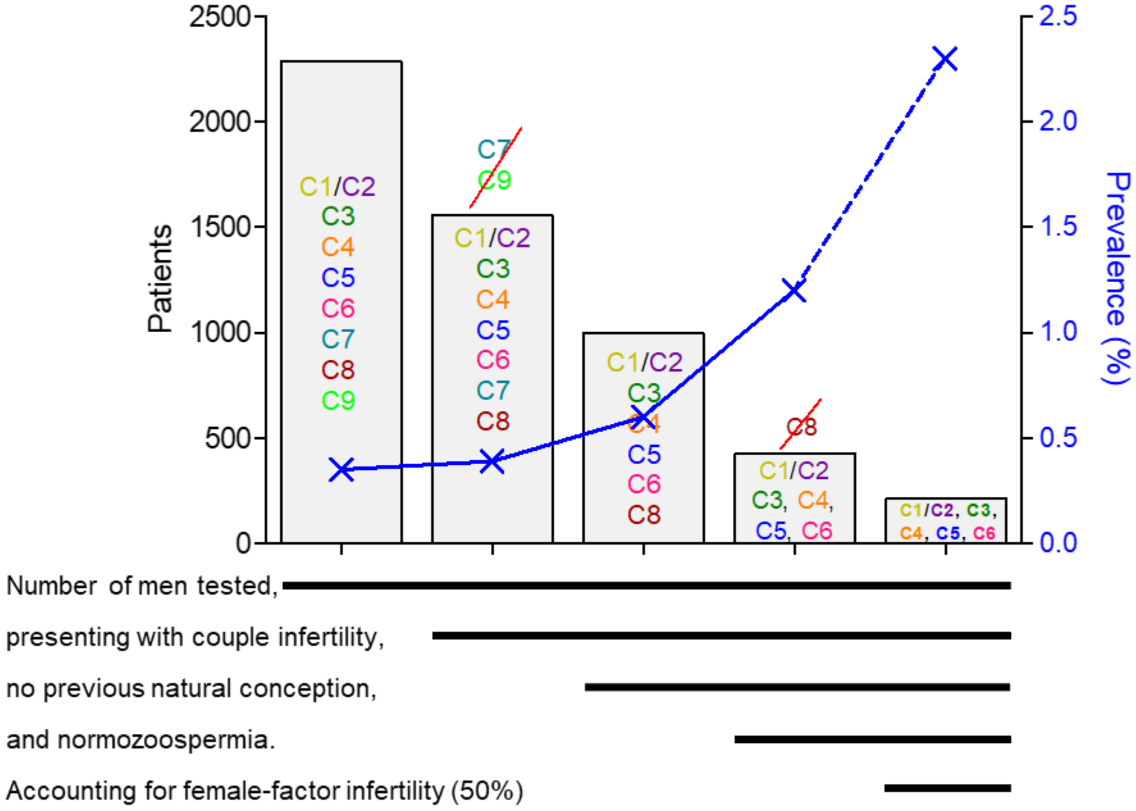


**Supplementary Figure 4: Stratification of men enrolled in the study and estimation of prevalences for CatSper-related male-factor infertility.** Among a total of 2,286 patients enrolled, eight patients with loss (C1-C8) and one (C9) with severely impaired CatSper function were identified. Among them were two brothers (C1 and C2) counting as one for the calculation of prevalence (blue). Of these, 1,557 patients, but not patient C7 and C9, presented because of couple infertility, 997 of which with no previous natural conception, of which 426 were also normozoospermic (excluding patient C8). Assuming that for about 50% of these men/couples the infertility is rather due to a female factor, we can interpolate a prevalence of 2.3% for a CatSper-related male-factor infertility (gray box / dashed line) among couples presenting with unexplained infertility.

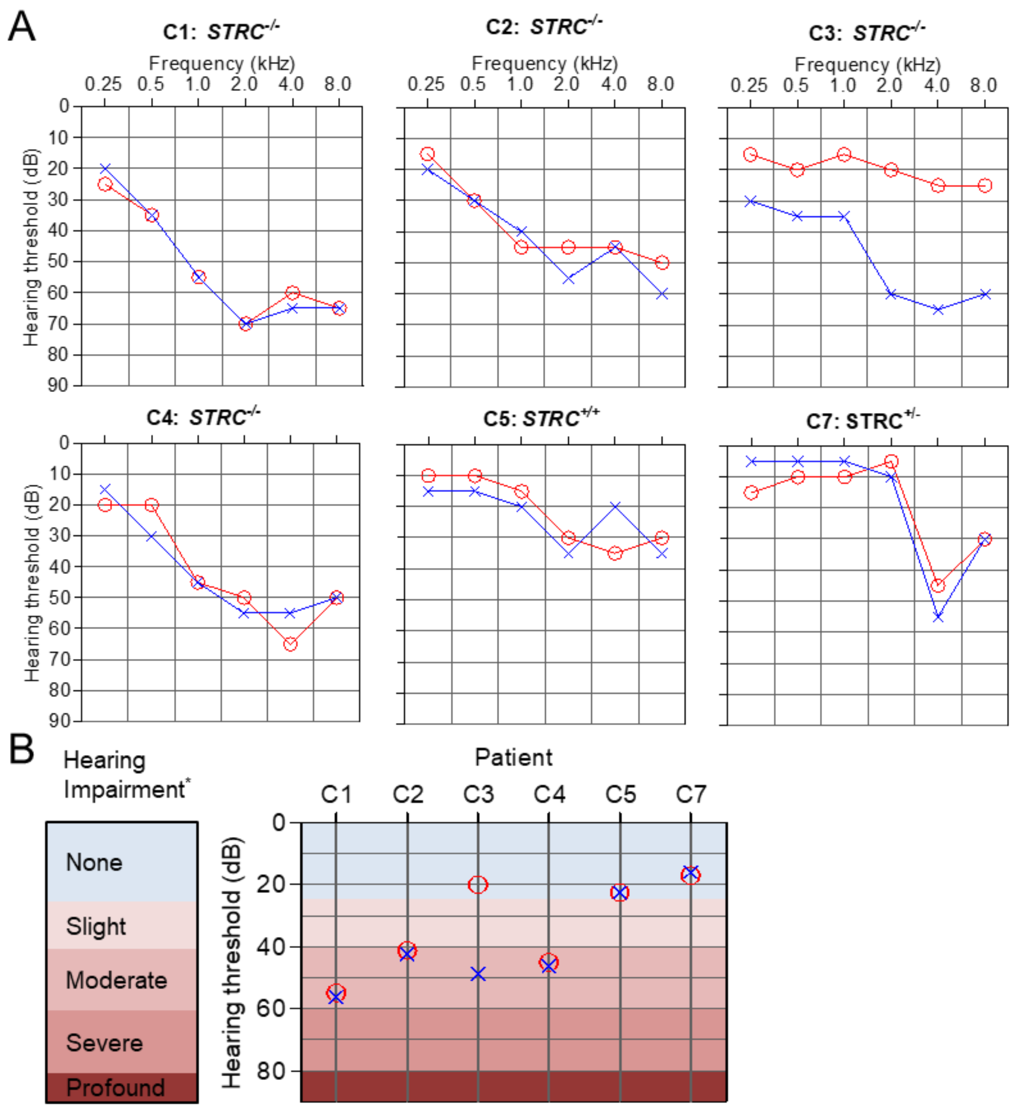


**Supplementary Figure 5: Audiometric analysis of six *CATSPER2*$^{-/-}$ patients: (A)** Audiograms depicting the hearing threshold of a sound at a given frequency in the right (red circles) and left ear (blue crosses). *STRC*$^{-/-}$ patients (i.e. patients C1, C2, C3, and C4) characteristically had sloping, high-frequency hearing impairment. **(B)** Summary of the audiometric analysis and scoring of the hearing impairments. Loss of *STRC* translated into moderate hearing loss, except in the case of patient C3, who suffered from moderate, unilateral hearing loss. Patient C7 that was heterozygous for *STRC* (*STRC*$^{+/-}$) and patient C5 with unaffected *STRC* presented with normal hearing. Hearing impairment is defined by the WHO as the averaged hearing threshold at 500, 1000, 2000, and 4000 Hz.

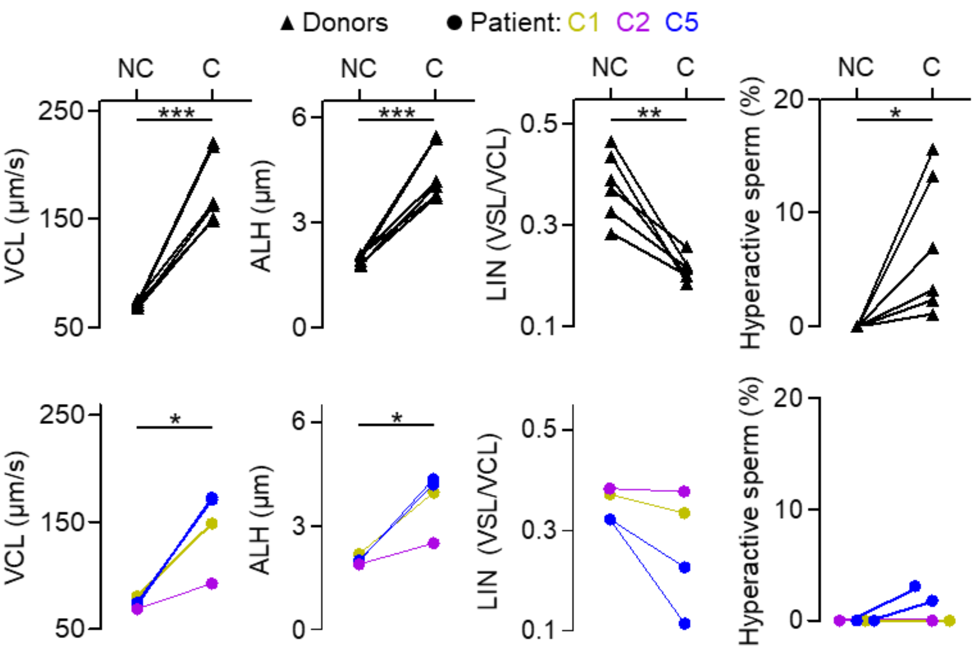


**Supplementary Figure 6: Spontaneous hyperactivation of control and *CATSPER2*$^{-/-}$ sperm.** Paired plots of kinematic parameters and fraction of hyperactivated sperm from donors (black triangles, n = 6) and *CATSPER2*$^{-/-}$ patients (color-coded circles, n = 4, i.e., four independent experiments with sperm from patients C1, C2, C5) incubated in parallel for at least three hours under non-capacitating (NC) and capacitating (C) conditions, respectively. These paired comparisons indicate that both in donors and *CATSPER2*$^{-/-}$ patients, capacitation affected CASA parameters when averaged over the entire sperm population. However, on the single sperm level, only in donors but not in *CATSPER2*$^{-/-}$ patients, a significant fraction of sperm exceeds the hyperactive-motility threshold upon capacitation. *P < 0.05, **P < 0.01***P < 0.001, paired t-test.

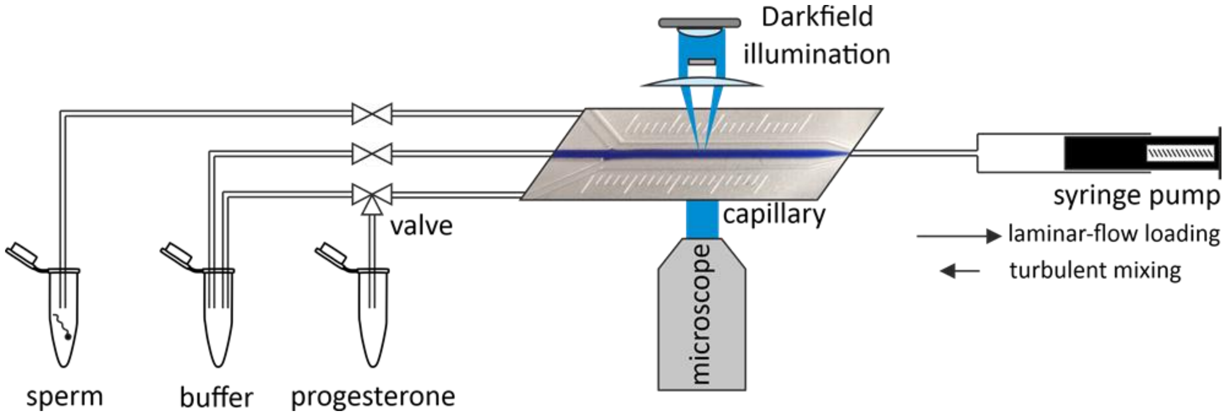


**Supplementary Figure 7: Kinetic CASA.** Illustration of the microfluidic set-up allowing for rapid mixing of sperm with progesterone and time-resolved analysis of ensuing motility responses by CASA. In brief, parallel laminar flows of two different solutions were established in a microcapillary. Using a syringe pump, sperm in $HTF^{++}$ were pulled through the capillary along with HTF++ containing progesterone or $HTF^{++}$ alone. Upon brief reversal of the flow direction, the solutions were rapidly mixed and the flow was stopped. Subsequently, every 15 seconds, the fraction of hyperactive sperm was determined by CASA and corrected for the basal level determined upon mixing of sperm with $HTF^{++}$ alone.

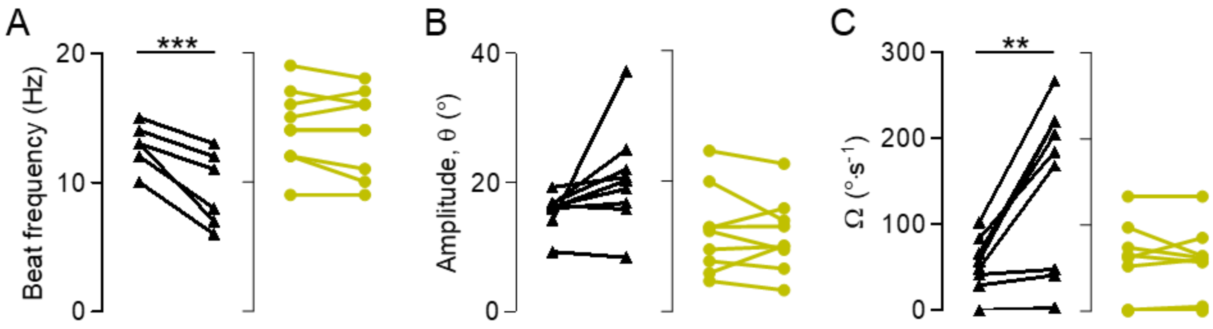


**Supplementary Figure 8: Frequency and amplitude of the flagellar beat and the rotation velocity of control sperm from donors and *CATSPER2*$^{-/-}$ sperm from patient C1 (see Figure 6A, B). (A-C)** Paired plots with mean ± SD (upper panels) and bar graphs (mean ± SD) (bottom panels) depicting the beat frequency (A), beat amplitude, θ (B), and rotation velocity Ω(°·s$^{-1}$) (C) of control (black, n = 9 from two donors) and patient C1 (gold, n = 9) sperm before and after uncaging progesterone. **P < 0.01, ***P < 0.001, paired t-test.

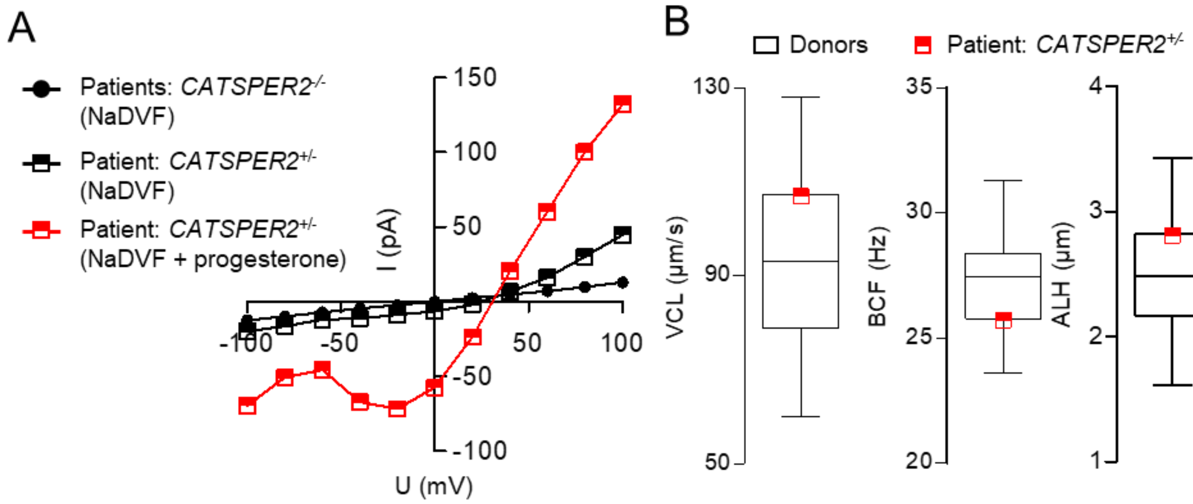


**Supplementary Figure 9: Patch-clamp recordings and basal-motility analysis of sperm from a *CATSPER2*$^{+/-}$ patient. (A)** Current-voltage relationship of monovalent currents in a sperm cell from a *CATSPER2*$^{+/-}$ patient perfused with divalent-free $Na^+$-based solution (NaDVF) (half-filled black squares) and NaDVF fortified with progesterone (2 µM) (half-filled red squares), and monovalent currents in sperm cells from *CATSPER2*$^{-/-}$ patients perfused with NaDVF (black circles) (mean ± SD, n = 5). The membrane voltage was stepped from -100 to +100 in increments of 10 mV from a holding potential of -80 mV. **(B)** Box and whiskers plots (whiskers: min to max) of kinematic parameters of control sperm from donors depicted in Fig. 4 (black, n = 23) and from a *CATSPER*$^{+/-}$ patient (half-filled red squares).

**Supplementary Table S1. CatSper channel subunits with the corresponding murine and human gene names.**

| **CatSper subunit (protein)** | **Gene name (murine)** | **Gene name (human)** |
|---|---|---|
| CatSper1 | *Catsper1* | *CATSPER1* |
| CatSper2 | *Catsper2* | *CATSPER2* |
| CatSper3 | *Catsper3* | *CATSPER3* |
| CatSper4 | *Catsper4* | *CATSPER4* |
| CatSperβ | *Catsperb* | *CATSPERB* |
| CatSperγ | *Catsperg* | *CATSPERG* |
| CatSperδ | *Catsperd* | *CATSPERD* |
| CatSperε | *Catspere* | *CATSPERE* |
| CatSperζ | *Catsperz* | *CATSPERZ* |
| CatSperη | *Catsperh* | *CATSPERH* |
| CatSperт | *Catspert* | *CATSPERT* |
| Efcab9 | *Efcab9* | *EFCAB9* |
| Slco6c1 | *Slco6c1* | *SLCO6A1* (ortholog) |
| Tmem249 | *Tmem249* | *TMEM249* |
| Trim69 | *Trim69* | *TRIM69* |

**Supplementary Table S2. Primer sequences used for Sanger validation of *CATSPERE* variants in patient C9.**

| **Primer** | **Sequence 5´-3´** |
|---|---|
| CATSPERE EX8_F | GCT GAA TCT TGA GCA CAT GGT AAA TTT |
| CATSPERE EX8_R | AGG ATT TGC CAC CAA CCT GA |
| CATSPERE EX18F | GCT GGA ACT CTT AAC CCC TCT |
| CATSPERE EX18R | TCC TAC CCA CTG CTG CCT TA |

**Supplementary Table S3. Results of SNP-array analysis.**

| Patient | Allel | Chromosome | Start | End | Size (bp) | *CATSPER2* deletion/zygosity |
|---|---|---|---|---|---|---|
| C1 | Allele 1 | 15q15.3 | 43,892,807 | 43,939,642 | 46,836 | homozygous |
| | Allele 2 | 15q15.3 | 43,892,807 | 43,939,642 | 46,836 | |
| C2 | Allele 1 | 15q15.3 | 43,892,807 | 43,939,642 | 46,836 | homozygous |
| | Allele 2 | 15q15.3 | 43,892,807 | 43,939,642 | 46,836 | |
| C3 | Allele 1 | 15q15.3 | 43,892,807 | 43,939,642 | 46,836 | homozygous |
| | Allele 2 | 15q15.3 | 43,892,807 | 43,939,642 | 46,836 | |
| C4 | Allele 1 | 15q15.3 | 43,892,807 | 43,939,642 | 46,836 | homozygous |
| | Allele 2 | 15q15.3 | 43,892,807 | 43,939,642 | 46,836 | |
| C5 | Allele 1 | 15q15.3 | 43,916,071 | 43,939,642 | 23,572 | homozygous |
| | Allele 2 | 15q15.3 | 43,916,071 | 43,939,642 | 23,572 | |
| C6 | Allele 1 | 15q15.3 | 43,916,071 | 43,939,642 | 23,572 | compound heterozygous |
| | Allele 2 | 15q15.3 | 43,892,807 | 43,939,642 | 46,836 | |
| C7 | Allele 1 | 15q15.3 | 43,916,071 | 43,939,642 | 23,572 | compound heterozygous |
| | Allele 2 | 15q15.3 | 43,892,807 | 43,939,642 | 46,836 | |
| C8 | Allele 1 | 15q15.3 | 43,916,071 | 43,939,642 | 23,572 | compound heterozygous |
| | Allele 2 | 15q15.3 | 43,892,807 | 43,939,642 | 46,836 | |
| C9 | no deletion | | | | | |

**Supplementary Movie S1**

Representative video of the ejaculate of a *CATSPER2*$^{-/-}$ patient

**Supplementary Movie S2**

Video of an IVF attempt with sperm from the *CATSPER2*$^{-/-}$ patient C5

**Supplementary Movie S3**

Video of head-tethered control sperm before and after uncaging progesterone

**Supplementary Movie S4**

Video of head-tethered *CATSPER2*$^{-/-}$ sperm before and after uncaging progesterone
